# Association of *ESR1* germline variants with *TP53* somatic variants in breast tumors in a genome-wide study

**DOI:** 10.1101/2023.12.06.23299442

**Authors:** Nijole P. Tjader, Abigail J. Beer, Johnny Ramroop, Mei-Chee Tai, Jie Ping, Tanish Gandhi, Cara Dauch, Susan L. Neuhausen, Elad Ziv, Nereida Sotelo, Shreya Ghanekar, Owen Meadows, Monica Paredes, Jessica Gillespie, Amber Aeilts, Heather Hampel, Wei Zheng, Guochong Jia, Qiang Hu, Lei Wei, Song Liu, Christine B. Ambrosone, Julie R. Palmer, John D. Carpten, Song Yao, Patrick Stevens, Weang-Kee Ho, Jia Wern Pan, Paolo Fadda, Dezheng Huo, Soo-Hwang Teo, Joseph Paul McElroy, Amanda Ewart Toland

## Abstract

**Background:** In breast tumors, somatic mutation frequencies in *TP53* and *PIK3CA* vary by tumor subtype and ancestry. HER2 positive and triple negative breast cancers (TNBC) have a higher frequency of *TP53* somatic mutations than other subtypes. *PIK3CA* mutations are more frequently observed in hormone receptor positive tumors. Emerging data suggest tumor mutation status is associated with germline variants and genetic ancestry. We aimed to identify germline variants that are associated with somatic *TP53* or *PIK3CA* mutation status in breast tumors.

**Methods:** A genome-wide association study was conducted using breast cancer mutation status of *TP53* and *PIK3CA* and functional mutation categories including *TP53* gain of function (GOF) and loss of function mutations and *PIK3CA* activating/hotspot mutations. The discovery analysis consisted of 2850 European ancestry women from three datasets. Germline variants showing evidence of association with somatic mutations were selected for validation analyses based on predicted function, allele frequency, and proximity to known cancer genes or risk loci. Candidate variants were assessed for association with mutation status in a multi-ancestry validation study, a Malaysian study, and a study of African American/Black women with TNBC.

**Results:** The discovery Germline x Mutation (GxM) association study found five variants associated with one or more *TP53* phenotypes with *P* values <1×10^-6^, 33 variants associated with one or more *TP53* phenotypes with *P* values <1×10^-5^, and 44 variants associated with one or more *PIK3CA* phenotypes with *P* values <1×10^-5^. In the multi-ancestry and Malaysian validation studies, germline *ESR1* locus variant, rs9383938, was associated with the presence of *TP53* mutations overall (*P* values 6.8×10^-5^ and 9.8×10^-8^, respectively) and *TP53* GOF mutations (*P* value 8.4×10^-6^). Multiple variants showed suggestive evidence of association with *PIK3CA* mutation status in the validation studies, but none were significant after correction for multiple comparisons.

**Conclusions:** We found evidence that germline variants were associated with *TP53* and *PIK3CA* mutation status in breast cancers. Variants near the estrogen receptor alpha gene, *ESR1,* were significantly associated with overall *TP53* mutations and GOF mutations. Larger multi-ancestry studies are needed to confirm these findings and determine if these variants contribute to ancestry-specific differences in mutation frequency.

## BACKGROUND

*TP53* and *PIK3CA* are among the most frequently mutated genes in breast tumors [1]. The frequency of somatic mutations in these genes varies by tumor subtype as well as ancestry [1–4]. Pan-cancer and breast cancer specific studies have found that tumors arising in individuals of African ancestry (AFA), particularly West African ancestry, are more likely to have somatic *TP53* mutations and less likely to have somatic *PIK3CA* mutations than tumors arising in individuals of European ancestry (EUR) [2–9]. *TP53* somatic mutations are more common in triple negative [estrogen receptor (ER) negative, progesterone receptor (PR) negative, human epidermal growth factor receptor 2 (HER2) negative] breast cancers (TNBC), while *PIK3CA* mutations are more common in hormone receptor (HR) positive HER2-tumors. However, even after adjusting for breast cancer subtype, ancestral differences in *TP53* and *PIK3CA* somatic mutation frequencies persist for some subtypes [2–4,10]. For example, one study found that 39% of hormone receptor (HR) positive HER2-tumors from individuals of AFA had *TP53* alterations compared to 24% of those of EUR [8]. Similarly, in HR+ HER2-tumors *PIK3CA* somatic mutations are less frequent in individuals of AFA (26%) versus EUR (42%) [8]. The biological mechanisms leading to the observed differences in *TP53* and *PIK3CA* somatic mutation frequency across populations and breast tumor subtypes are not understood.

*TP53* encodes transcription factor TP53 and is mutated in a high proportion of breast and other cancers, resulting in altered expression of genes important for response to cellular stress and apoptosis. Unlike many genes involved in tumorigenesis, *TP53* can have either loss of function (LOF) mutations, which lead to total loss of the ability of the protein to transactivate, or gain of function (GOF) mutations, which result in TP53 binding to new promoters to activate genes not typically associated with TP53 [11,12].

TP53 is a tetramer but can also bind to related proteins TP63 and TP73 [13]. Some TP53 tumor-associated mutations act in a dominant negative manner where the mutant version of the protein interferes with the function of wildtype proteins in the tetramer. In previous studies, we found that in breast tumors with *TP53* mutations, those from AFA women were less likely to have GOF mutations than those from EUR women [14]. Mutations without dominant negative activity were associated with TNBC and ER negative status. These data suggest that types of *TP53* mutations in breast tumors differ by self-reported race and tumor subtypes which may be due to different functional consequences of these mutations within cells.

Whilst most somatic events in tumors are likely due to exogenous or endogenous mutators, recent evidence suggests that germline variants may influence the type and burden of somatic changes. Tumor mutational burden, caused in part by somatic mutations in DNA repair genes, is a polygenic trait with an estimated 13% of the variation explained by common germline variants [15]. Some tumor mutational signatures are associated with common inherited variants in genes such the apolipoprotein B mRNA editing enzyme catalytic polypeptide (APOBEC) mutation signature and variants in *GNB5* [16].

Pathogenic variants (PV) in high-risk cancer susceptibility genes also associate with the presence of somatic mutations and specific mutational signatures. Breast tumors arising in individuals with a germline *BRCA1* PV have more frequent occurrence of somatic *TP53* mutations compared to those without a *BRCA1* PV [17–20]. Breast and ovarian tumors arising in individuals with *BRCA1* and *BRCA2* PVs typically show homology directed repair deficiency signatures [20,21].

Based on these studies, we hypothesized that the germline genetic background of an individual can influence specific mutational processes, tumor promotion, and/or mutations in specific cancer-related genes during tumorigenesis, any of which could lead to the observed differences in the frequency of key cancer driver mutations by ancestry [22]. The goal of this study was to identify inherited common germline variants (G) that are associated with *TP53* or *PIK3CA* somatic mutation status (M) in breast tumors using a **G**ermline Variant by **M**utation (GxM) genome-wide association study (GWAS) design to assess the influence of genetic background on mutation frequency of these genes.

## METHODS

### Discovery Breast Cancer Datasets

Existing datasets of women with breast cancer from the Cancer Genome Atlas (TCGA) [1], Molecular Taxonomy of Breast Cancer International Consortium (METABRIC) [23], and the Welcome Trust Sanger Institute [24] were used for the discovery GxM GWAS. Each study had existing genome-level single nucleotide variant (SNV) genotyping data, somatic mutation data for *TP53* and *PIK3CA*, and associated clinical and tumor details such as self-reported race/ethnicity, age at diagnosis (reported as decade of diagnosis), and ER, PR and HER2 status (Supplemental Information Tables 2-4).

### *PIK3CA* and *TP53* Somatic Mutation Classification

For discovery and validation analyses, *PIK3CA* mutation status (yes or no) was defined for the following phenotypes: any non-loss of function somatic variant in *PIK3CA*, any activating/hot-spot mutation [25], or specific activating mutations (e.g. p.E542K, p.E545K, p.H1047R/L). *TP53* mutation status was classified as the presence of any somatic variant in a codon exon or splice-site (yes/no), and variants resulting in *TP53* LOF or GOF as described (Supplemental Information Table S5) [14,26]. Somatic variants that resulted in a synonymous change and were not predicted to affect splicing were not considered to be a mutation. GOF mutations displayed one or more of the following phenotypes in functional studies: interference with TP63 or TP73 activity, transactivation of genes repressed by wildtype TP53, or cooperation with oncogenes in rat or mouse embryonic fibroblasts [27–61]. *TP53* LOF variants were those that abolished transactivation activity and/or resulted in altered splicing, frameshift, or nonsense changes. *TP53* somatic missense variants with insufficient data to functionally score as LOF or GOF were called unknown and were not included in LOF or GOF specific analyses (Supplemental Information Table S5). Larger copy number loss of *TP53* was not included as a mutation type in the analyses due to lack of annotated data for multiple datasets. Controls for each analysis were individuals with breast cancer with no somatic mutation in the gene being assessed.

### Ancestry and GWAS analyses

PLINK was used to merge data sets, filter, and analyze data. Ancestry SNVs for principal component analyses (PCA) were determined using the Affymetrix annotation accomplished by subtracting the minor allele frequency (MAF) from each of four populations [Han Chinese in Beijing (CHB), Yoruba in Ibadan (YRI), Northern Europeans from Utah (CEU), Japanese in Tokyo (JPT)] in the annotation in a pairwise manner and taking the top 1000 SNVs from each comparison. This resulted in the use of 4486 unique “ancestry” SNVs; 4212 of those had a MAF of greater than 1%. PCA were performed on these 4212 SNVs to identify individuals of non-European ancestry (e.g. those who did not cluster with the European ancestry group); these individuals were removed from the discovery analyses and were included in the validation studies. There was a high concordance of ancestry assignment with self-reported race. Filtering also included removal of SNVs with MAF less than 0.01, male participants, and samples and SNVs with greater than 10% missing values. Imputed SNVs were not included. SNVs showing Hardy Weinberg equilibrium *P* values less than 1×10^-50^ were also removed. Association analyses were run on a final set of 2850 females of European ancestry and 739,537 SNVs with PLINK using a logistic model with a covariate for study. An additive model was assumed. *P* values were false discovery rate (FDR) corrected and visualized using R [62].

### Selection of variants for validation analyses

Variants were prioritized for validation studies through multiple qualitative and quantitative filtering steps (Supplemental Information Tables S6 and S7). Information used to rank variants included *P* values <1×10^-4^, odds ratios (OR), MAF greater than 10% in at least one of three populations (European, African or East Asian), allele frequency differences by ancestry, proximity to a variant identified in GWAS for breast cancer risk or other relevant phenotypes (e.g. other cancers, age at menarche, obesity), proximity to a gene showing a role in tumorigenesis, and mapping to a functionally active region (e.g. transcription start site, active chromatin markers, estimated or actual transcription factor binding site, disruption of a transcription factor binding motif, ChIP-seq region for breast cancer cell line, characterized gene enhancer, characterized promoter region, or expression quantitative trait locus).

Online resources used for *in silico* screening of candidate SNVs included UCSC Genome Browser (https://genome.ucsc.edu/), GTEx Portal (https://gtexportal.org/home/), RegulomeDB v.2.0.3 (https://www.regulomedb.org/regulome-search/), HaploReg v4.0 (https://pubs.broadinstitute.org/mammals/haploreg/haploreg.php), and dbSNP (https://www.ncbi.nlm.nih.gov/snp/). In addition to variants chosen from the discovery GxM GWAS findings, additional variants were analyzed including two SNVs mapping near *SETD9/MAP3K1* previously shown to be associated with *PIK3CA* somatic mutations in breast cancer [63], an *XPC* variant rs2228001 previously shown to be associated with *TP53* mutation status [64], and a variant in *AURKA* (rs2273535) shown in our unpublished studies to be associated with somatic GOF *TP53* mutations in human papilloma virus negative head and neck squamous cell carcinomas from TCGA. When a genotyping assay for a variant could not be designed for technological reasons, another variant from that locus or a variant in high linkage disequilibrium (LD) (r^2^>0.8) with the original variant was included as a replacement.

### Validation Genotyping

Validation genotyping for 188 SNVs of interest (95 for *TP53* and 93 for *PIK3CA*) was completed for cohorts without existing genome-wide genotyping data including individuals from the Stefanie Spielman Breast Cancer Cohort (n=144), OSU Total Cancer Care (TCC) (n=351) and the City of Hope (COH) Latina Breast Cancer Study (n=120) using a Fluidigm HD Biomark in a 96x96 format in the Ohio State University (OSU) Comprehensive Cancer Center (CCC) Genomics Shared Resource (GSR) (Supplemental Tables S8, S9). Each genotyping plate contained two duplicate DNA samples, three no-template controls (water), and one control DNA sample genotyped on all plates. DNAs that failed for more than 10% of SNVs from a plate were repeated and if failed again were removed from analysis.

SNVs that failed for more than 10% of samples or failed to consistently form three clear genotyping groups were removed from analyses. For genetic ancestry, 96 SNVs were chosen for genotyping from existing ancestry informative marker (AIM) panels [65–67] (supplemental table S10-S12). Of the 96 AIM SNVs, two were removed for poor genotyping performance.

### Somatic Mutational Analyses

For the validation studies, *TP53* and *PIK3CA* mutational status from the clinical testing reports or targeted or exome sequencing of tumor DNA was available for breast cancer cases from the COH Latina Breast Cancer Study, TCGA, and a subset of the TCC cases. For cases in which mutation status was not known, tumor tissue or DNA was available from the Spielman Breast Cancer Cohort and the TCC program.

#### Next-generation sequencing mutational analysis

An Agilent custom-targeted sequencing panel that included *TP53* and *PIK3CA* was designed for an unrelated project. Tumor DNA samples isolated from frozen tumors from TCC cases (n=12) were assessed for somatic mutations in *TP53* and *PIK3CA* using this panel. Libraries were generated according to Agilent protocols and sequenced on a HiSeq4000 by the OSU CCC GSR. Sequencing run data was received in the form of paired FASTQ files (forward and reverse reads). FASTQ file pairs were aligned to the hg19 reference genome using Genomic short-read nucleotide alignment program (GSNAP) (version 2019-03-04)[68]. Variants were called with Pisces (version 5.2.10.49) using the hg19 reference genome [69]. At this stage, low depth (<100 read count) variants were removed. Finally, both SnpSift (version 4.3t) and Annovar (version 20190316) were used to annotate called variants [70,71].

#### Sanger sequencing mutational analysis

The remainder of tumor samples lacking existing somatic mutation data (n=126 for *TP53*, n=184 for *PIK3CA*) were screened for somatic mutations in *TP53* coding exons (exons 2-10) and *PIK3CA* exons 4, 9 and 20 using Sanger sequencing. Tumor DNA (10-20ng) was PCR amplified and products were confirmed for size by gel electrophoresis (Supplemental Table S13). PCR products were Exo/SAP-IT treated and Sanger sequenced in both forward and reverse directions by the GSR. Sequence chromatograms were evaluated for mutations using DNASTAR Lasergene v.17 by two different laboratory members.

### GxM Validation Analyses

Data used for validation of key findings included genotype and tumor mutational data from individuals of non-European ancestry from the three discovery datasets as well as samples (germline and/or tumor DNA) or existing data from 1285 individuals of multiple ancestries from the METABRIC (n=166), Stefanie Spielman Breast Cancer Cohort (n=144), OSU TCC (n=352), a Nigerian breast cancer study (n=100), the COH Latina Breast Cancer Study (n=120), TCGA (n=302), and a TCGA study (“Banerji study”) of women from Mexico and Vietnam (n=101) (Supplemental Tables S2-S4, S14-S18)[3,4,72,73]. Genetic ancestry by PCA classified 341 women as AFA (26.5%), 572 women as EUR (44.5%), and 133 women as East Asian ancestry (EAS) (10.4%). The remainder of women (18.6%) were admixed (falling between PC clusters), most of whom self-identified as Hispanic/Latino. Due to some missing genotypes, not every variant had data for all 1285 individuals (Supplemental Table S26-S27).

For association analyses, logistic models were employed with an additive effect for the SNV. Study and ancestry were included as covariates in the models. For the study and ancestry-specific analyses, the study analysis omitted the effect of study, and the ancestry analyses omitted the ancestry principal component (PC) from the models. Because two different panels were used for ancestry determination, individuals of known ancestry (HapMap, TCGA) were used as anchors for each panel. The PC1/PC2 were rotated so that the known ancestry groups overlapped and the distance from the anchor group was calculated as the PC covariables. For individuals with available genome-wide genotyping data, imputation of validation SNVs not present on the GWAS genotyping panels was performed. Imputation was carried out after removing genotypes with no calls or Y chromosome calls. Eagle was used to phase SNV, and imputation was done using Minimac3. The maximum expected error rate across imputed validation SNPs was 0.086. Formats were converted to PLINK format, and variants with greater than 2 alleles were removed.

### Independent Validation Studies

SNVs of interest also were assessed independently in two cohorts: 859 women with breast cancer from the Malaysian Breast Cancer Study (MyBrCa) [74,75] and 393 AFA women with TNBC from the Breast Cancer in African Americans: Understanding Somatic Mutations and Etiology (B-CAUSE) study (Supplemental Tables S19 and S20) [76]. Validation SNVs for the MyBrCa study were excluded from analyses if they mapped to the X chromosome or if they had a MAF less than 1% in Malaysian individuals. SNVs were excluded from analyses for the B-CAUSE study if they mapped to the X chromosome. For the MyBrCa study association tests were conducted using SNPtest adjusted to information for ancestry [4 PCs], age of diagnosis, and ER status. B-CAUSE data came from women who self-identified as Black and were diagnosed with TNBC. The African-ancestry Breast Cancer Genetic (AABCG) is a large breast cancer consortium which provided genome-wide genotyping data for the B-CAUSE study. AFA ancestry was confirmed by estimating global African ancestry using ADMIXTURE [77] (Supplemental Tables S20). As the frequency of somatic TP53 mutations in the B-CAUSE TNBC cases was high, analyses were run for *TP53* GOF-associated germline variants using individuals with LOF *TP53* mutations and those with no mutations as controls; conversely for *TP53* LOF-associated variants, analyses were run using individuals with GOF plus those with no mutations as controls.

## RESULTS

To identify germline variants associated with *TP53* or *PIK3CA* somatic mutations in tumors, we identified existing datasets with GWAS-level germline variant information, somatic mutation information for *TP53* and *PIK3CA*, and demographic and clinical information such as age of diagnosis, tumor subtype defined by hormonal (ER and PR) status and HER2 amplification. Three datasets were identified that fit these criteria (Supplemental Tables S2-S4). After filtering for SNVs with MAF less than 1%, individuals with 10% or higher SNV genotypes missing, SNVs out of Hardy-Weinberg equilibrium (p<1×10^-50^) and individuals of non-European ancestry, 2850 females of EUR with breast cancer and 739,537 SNVs were included in the discovery GWAS for variants associated with *TP53* and *PIK3CA* mutation status.

### Discovery GxM for *TP53* mutation status

Somatic mutations in *TP53* (237 GOF, 536 LOF, 106 unknown) were identified in 879 of the 2850 women (30.8%) (Supplemental Tables S2-S5). Analyses for association with any *TP53* mutation, GOF *TP53* mutation, and LOF *TP53* mutation were performed. Following analysis, no SNV met the genome-wide statistical significance threshold of a *P* value <1.0×10^-8^; four variants were identified with *P* values ≤1.0×10^-6^ and 34 variants had *P* values less than ≤1.0×10^-5^ across 22 loci (Figure 1A, 1B and 1C; Supplemental Table S21 and S22). Two variants showed *P* values of <1.0 x 10^-5^ for more than one *TP53* phenotypic comparison: rs1561072 for any *TP53* mutation and GOF *TP53* mutations and rs2886631 for any *TP53* mutation and LOF *TP53* mutations.

**Figure 1:**
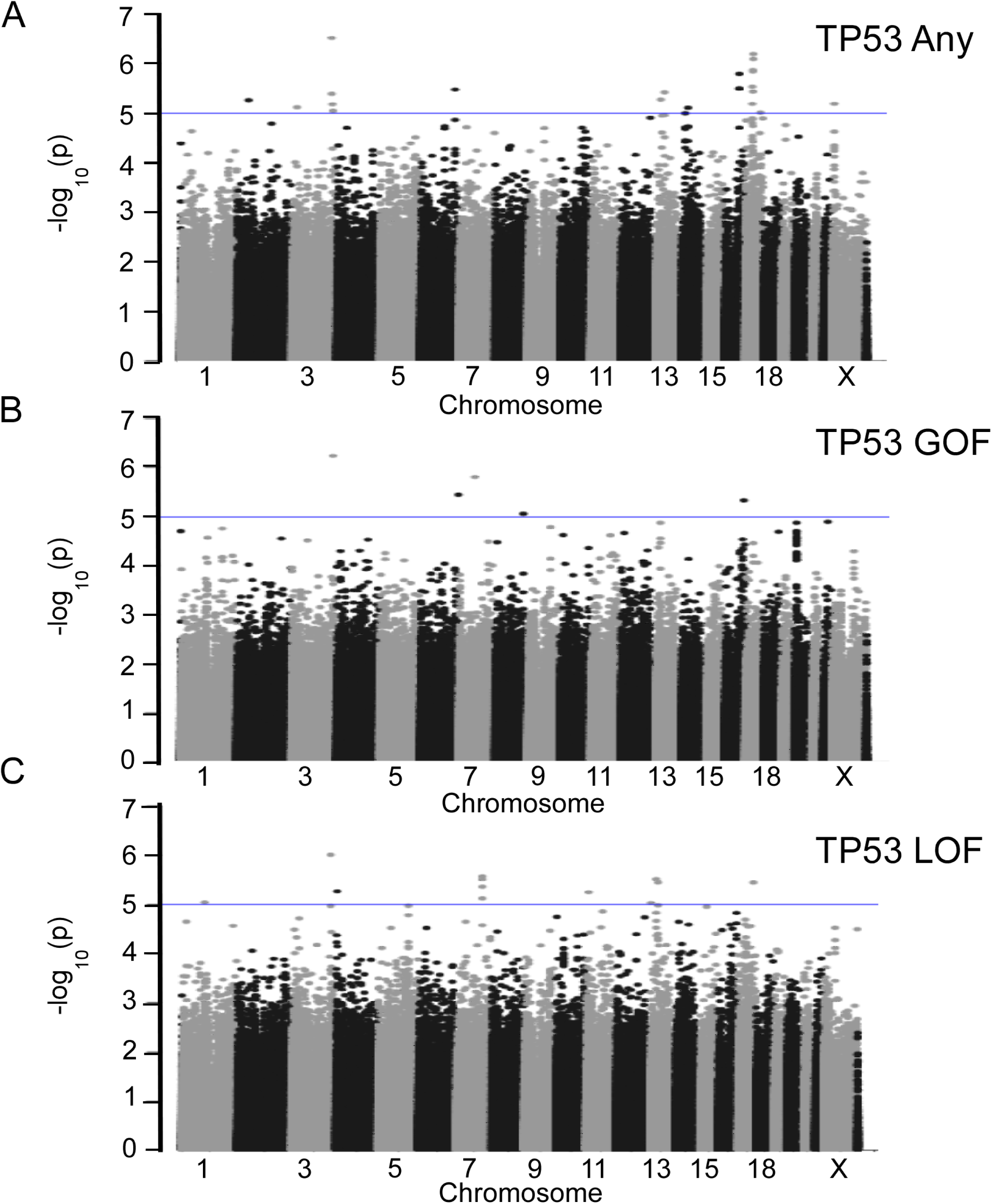
Manhattan Plots for *TP53* Discovery GxM Analyses. Discovery GWAS data for (A) 879 *TP53* mutation carriers and 1965 breast cancer cases without *TP53* mutations, (B) 237 cases with *TP53* GOF mutations and 1965 breast cancer cases without *TP53* mutations, and (C) 536 cases with *TP53* LOF mutations and 1965 controls are plotted by -log_10_(*P* values). Blue lines represent *P* values of less than 1×10^-5^. Chromosome numbers are indicated. GXM, Germline Variant by Mutation; GWAS, genome-wide association study; GOF, gain of function; LOF, loss of function.

**Figure 2:**
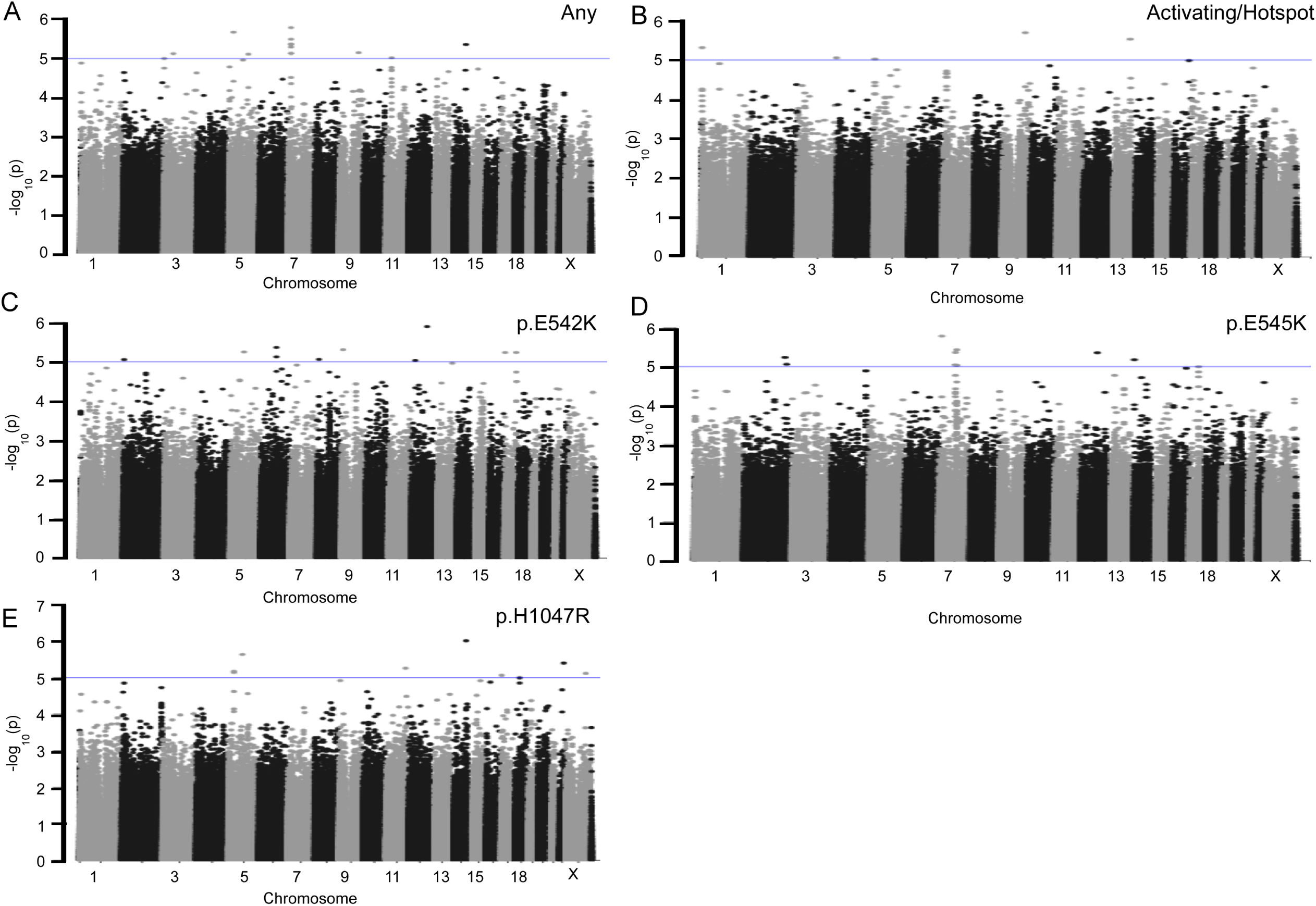
Manhattan Plots for *PIK3CA* Discovery GxM Analyses. Discovery GWAS data for (A) 1095 *PIK3CA* mutation carriers and 1642 breast cancer cases without *PIK3CA* mutations, (B) 858 cases with PIK3CA activating/hotspot mutations and 1642 breast cancer cases without *PIK3CA* mutations, (C) 112 cases with *PIK3CA* p.E542K mutations and 1642 breast cancer cases without *PIK3CA* mutations, (D) 193 cases with *PIK3CA* p.E545K mutations and 1642 breast cancer cases without *PIK3CA* mutations, and (E) 387 cases with *PIK3CA* p.H1047R mutations and 1642 breast cancer cases without *PIK3CA* mutations are plotted by -log_10_(*P* value). Blue lines represent *P* values of less than 1×10^-5^. Chromosome numbers are indicated. GXM, Germline Variant by Mutation; GWAS, genome-wide association study.

### Discovery GxM for *PIK3CA* mutation status

Thirty-eight percent (n=1095) of breast tumors from the 2850 women included in the discovery analyses had a somatic *PIK3CA* mutation. Of these, 858 tumors had a mutation known to functionally activate *PIK3CA* encoded protein PIK, including 112 with p.E542K, 193 with p.E545K, and 387 with p.H1047R/L alterations. Following association analyses for *PIK3CA* mutation status, no SNV met genome-wide significance of *P* value of <1.0×10^-8^ (Figure 1B). Forty-four SNVs were associated with one or more *PIK3CA* mutation phenotypes with *P* value <1×10^-5^ (Figure 1B; Supplemental Table S23 and S24). Of these, rs2026801 showed evidence of association (*P* value <1×10^-5^) for any *PIK3CA* mutation and activating *PIK3CA* mutations, and rs1712829 showed evidence of association with both p.H1047R and any *PIK3CA* mutation.

### Selection of variants for validation studies

Using *in silico* filtering approaches, all variants with *P* values < 1×10^-4^ for any phenotype were evaluated for potential inclusion in validation studies. Variants were prioritized for further evaluation by allele frequency (MAF>10%), potential function using *in silico* prediction models, location near a known GWAS hit for breast cancer or related phenotype (e.g. age of menarche, obesity), location near a gene involved in tumor development, or known relationship to PI3K or TP53 pathways (Supplemental Tables S6 and S17). Of these, 188 variants from *TP53* (n=95) and *PIK3CA* (n=93) GxM analyses were chosen for validation studies and successfully genotyped in multi-ancestral populations (Supplemental Table S25). For individuals with GWAS-level genotyping data, 119 variants for *TP53* and 106 variants for *PIK3CA* were tested (Supplemental Tables S26-S31).

### Mutation Status and Ancestry in Validation Populations

In the multi-ancestry validation data set, ancestry classifications by PCA yielded 340 AFA individuals, 602 EUR individuals and 134 EAS individuals. The remainder of study individuals (n=209) were considered admixed and not assigned to a specific group; these included individuals of Hispanic/Latino background who demonstrated a high degree of admixture. In the multi-ancestry validation cohort, 419 of 1036 individuals (40%) had a *TP53* somatic mutation of which 110 (10.6%) were GOF and 277 (27%) were LOF. The overall frequency of *PIK3CA* mutations in the multi-ancestry validation cohort was 28% (290 of 1036) with 235 of those being activating (81% of all mutations and 23% of the total). For specific *PIK3CA* activating mutations, 40/1035 (3.9%) of tumors had a p.E542K mutation, 58/1035 (5.6%) had a p.E545K mutation, and 133/1035 (13%) had a p.H1047R/L mutation.

The MyBrCa study included 859 women from Malaysia with breast cancer, of whom 369 carried a *TP53* mutation (43%). Of the MyBrCa women with a *TP53* mutation, 114 had a GOF mutation (13%), 241 (28%) had LOF mutations, and the remainder (n=14) were of unknown GOF/LOF status. For *PIK3CA* mutation status, 247 of the MyBrCa women (29%) had *PIK3CA* mutations in their breast tumor; of these, 217 (25% of total; 88% of all with a mutation), were activating. For the specific *PIK3CA* mutations, 20 had a p.E542K mutation (2% of total), 55 had a p.E545K mutation (6% of total) and 115 (13% of total) had a p.H047R/L mutation. Of the 393 women of African ancestry with TNBC in the B-CAUSE study, 365 (93%) had a TP53 somatic mutation including 260 (66%) with LOF mutations and 85 (22%) with GOF mutations. Only nine (2.3%) of tumors from women in the B-CAUSE study had any *PIK3CA* somatic mutations, of which four were activating/hotspot mutations.

### Association of variants at the *ESR1* locus and *TP53* mutation status

In the multi-ancestry validation study, rs9383938, which maps to chromosome 6 at the *ESR1* locus, was significantly associated with having any *TP53* mutation (OR 1.46, *P* value 6.8×10^-5^) after correction for multiple comparisons of 119 variants/*TP53* phenotypes (<0.00042). In the MyBrCa study, rs9383938 also showed association with having a *TP53* mutation (OR=1.81; *P* value 9.8×10^-8^) (Table 1) (Supplemental Tables S27 and S32) and *TP53* GOF mutation status (*P* value 8.4×10^-6^). Another *ESR1* locus variant, rs9479090, was associated with *TP53* mutations (*P* value 2.8×10^-7^) in the MyBrCa Study.

**Table 1:**
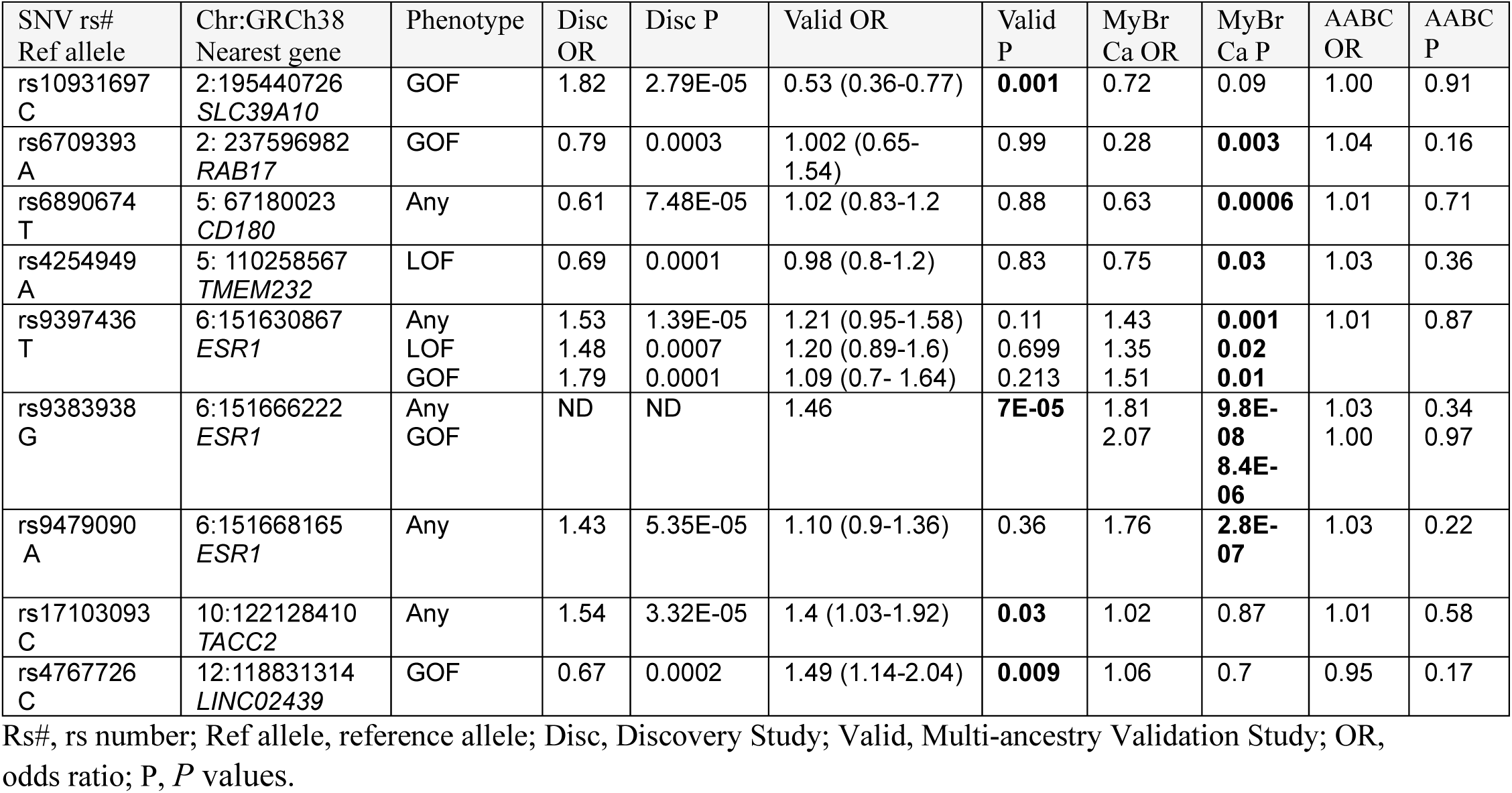
Select variants showing evidence of association with *TP53* mutation status.

After correcting for multiple comparisons, no other variants were significantly associated with any *TP53* phenotype in the validation analyses, MyBrCA, or B-CAUSE studies. However, multiple variants showed nominal *P* values (<0.05), including five in the multi-ancestry validation cohort (rs10931697, rs4767726, rs12470238, rs9911226, rs17103093), three in the MyBrCa Study (rs6890674, rs6709393, rs4254949), and four in the B-CAUSE study (rs7633912, rs9926714, rs28547342, rs12445424) (Table 1; Supplemental Tables S26-S28, S32-S33). Variants showing a trend for association in more than one dataset included rs10931697 in the multi-ancestry validation and MyBrCa studies (*P* values 0.001 and 0.09), and rs6709393 (*P* values 0.003 and 0.16) in the MyBrCa and B-CAUSE studies.

### Association of germline variants and *PIK3CA* mutation status

After correction for multiple comparisons for 106 variants/phenotype (*P* value <0.00047), no SNVs were significantly associated with any *PIK3CA* phenotype in the validation set, MyBrCA study (*P* value <0.00048) or B-CAUSE study (*P* values <0.0021) (Table 2) (Supplemental Tables S29-S31, S34-S35). Four variants showed nominal association (*P* value <0.05) for a *PIK3CA* mutation phenotype in the validation data set (rs6955337, rs10975835, rs13230836 and rs1886675) and nine variants in the MyBrCa study (rs2026801, rs1051894, rs8084310, rs2332431, rs479995, rs194537, rs859074, rs9791638, rs2127064) (Table 2). Three variants with *P* values for association with *PIK3CA* mutation status of less than 0.05 were observed in the B-CAUSE study (rs893344, rs252913, rs331499), but these data should be interpreted with caution due to the very small numbers of *PIK3CA* mutation carriers.

**Table 2:** Variants associated with *PIK3CA* mutation status.

| SNV Rs#<br>Ref Allele | Chr:GRCh38<br>Nearest Gene | Phenotype | Disc<br>OR | Disc P | Validation OR<br>(95% CI) | Validation<br>P | MyBrCa<br>OR | MyBrCa<br>P |
| --- | --- | --- | --- | --- | --- | --- | --- | --- |
| rs859074<br>G | 1:94883881<br>*SLC44A3* | E542K | 0.54 | 0.0002 | 1.02 (0.68-1.53) | 0.90 | 0.45 | **0.04** |
| rs13230836<br>A | 7:17974272<br>*SNX13/HDCA9* | Any | 0.78 | 7.41E-06 | 0.81 (0.68-0.97) | **0.023** | 1.2 | 0.12 |
| rs6955337<br>A | 7:17974951<br>*SNX13/HDCA9* | Any | 1.3 | 3.22E-06 | 1.41 (1.1-1.82) | **0.007** | 0.83 | 0.13 |
| rs9791638<br>T | 7:90158086<br>*STEAP1/STEAP2* | E545K | 1.68 | 1.54E-05 | 0.93 (0.59-1.47) | 0.78 | 1.67 | **0.047** |
| rs194537<br>T | 7:90245730<br>*STEAP1/STEAP2* | E545K | 1.73 | 4.6E-06 | 0.81 (0.54-1.19) | 0.53 | 1.67 | **0.042** |
| rs10975835<br>C | 9:6797498<br>*KDM4C* | H1047R/L | 1.54 | 1.12E-05 | 1.5 (1.1-2.04) | **0.011** | 0.92 | 0.66 |
| rs2026801<br>A | 9:111143171<br>*LPAR1* | Activating | 0.75 | 2.52E-06 | 0.88 (0.73-1.05) | 0.16 | 0.74 | **0.009** |
| rs2332431<br>A | 14:70609134<br>*MED6* | E542K | 1.80 | 0.0002 | 1.26 (0.80-1.93) | 0.31 | 0.46 | **0.03** |
| rs10518943<br>C | 15:57863151<br>*POLR2M*<br>*ALDH1A2* | E542K | 0.51 | 8.11E-05 | 0.99 (0.65-1.49) | 0.96 | 0.32 | **0.016** |
| rs8084310<br>T | 18:8950506<br>*SOGA2*<br>*CCDC165* | E542K | 1.80 | 0.0002 | 1.09 (0.63-1.82) | 0.75 | 3.39 | **0.017** |
| rs2442046<br>G | 2:135014078<br>*MGAT5* | E542K | 1.86 | 1.94E-05 | 1.64 (0.95-2.77) | 0.07 | 1.92 | 0.14 |
| rs2738348<br>C | X:348699 | E542K | 0.7 | 0.0003 | 0.78 (0.59-1.03) | 0.08 | ND | ND |

Ref allele, reference allele; CI, confidence interval; ND: No data; Disc, Discovery Study; Validation, Multi-ancestry Validation Study; OR, odds ratio; p, *P* values.

Variants showing a trend towards association in the multi-ancestry validation study and the MyBrCa study included rs2026801 with activating mutations (OR 0.88, *P* value 0.16 and OR 0.74, *P* value 0.009, respectively) and rs24420246 with p.E542K mutations (OR=1.64, *P* value 0.07 and OR=1.92, *P* value 0.14).

### Association with African Ancestry

As *TP53* mutation frequency is generally higher in breast tumors arising in individuals of AFA compared to other populations, we tested if any of the variants in our validation analyses showed stronger evidence of association in this population. Variants rs16951139, rs10514489 and rs10931697 showed nominal evidence of association with *TP53* GOF mutation status in AFA breast cancer cases from the multi-ancestry study (*P* values 0.018, 0.022 and 0.022, respectively) (Supplemental Table S28). rs10514489 also showed evidence of association with *TP53* GOF mutation status in individuals of EUR (*P* value 0.008) and rs10931697 showed a non-significant trend for association with GOF *TP53* status in the MyBrCa study (*P* value 0.09). In the B-CAUSE study, none of these variants showed evidence of association. However, four variants (rs7633912, rs9926714, rs28547342, and rs12445424) in the B-CAUSE study showed nominal evidence of association with one or more *TP53* phenotypes (*P* value < 0.05) (Supplemental Table S33).

*PIK3CA* mutation status and genotype data were available for 127 to 338 females of AFA in the multi-ancestry study depending on the variant. Three variants showing evidence of association with a *PIK3CA* phenotype in the discovery analysis showed nominal evidence of association in AFA women in the multi-ancestry study. These included rs2586532 (p.E542K, *P* value 0.005), rs4322362 (p.H1047R/L, *P* value 0.022) and rs3812471 (any *PIK3CA* mutation; *P* value 0.026) (Supplemental Table S31). None of these variants showed evidence of association in other populations or in the combined analysis. In the B-CAUSE study, rs893344 showed evidence of association for any *PIK3CA* somatic mutation (*P* value 0.044) and activating *PIK3CA* mutations (*P* value 0.09). While this variant did not show evidence of association in the multi-ancestry or MyBrCa studies, there was a trend towards association when only the AFA women in the multi-ancestry study were considered (*PIK3CA* any mutation *P* value 0.09 and activating mutation *P* value 0.13) (Supplemental Table S35).

## DISCUSSION

To our knowledge, this is the first genome-wide breast cancer specific study to identify germline variants that are associated with *TP53* or *PIK3CA* somatic mutation status. As different types of mutations may have differential effects on cancer-related phenotypes, we also tested for association of specific subcategories of *TP53* (any, LOF, GOF) and *PIK3CA* (any, activating, specific site) mutations with common SNVs. Five variants from the discovery analyses of women of EUR showed suggestive evidence (*P* value < 1×10^-6^) for association with *TP53* mutation status. Analyses of candidate variants in a multi-ancestry cohort and a Malaysian study, MyBrCa, confirmed that variants at the *ESR1* locus were associated with multiple *TP53* mutation classifications (any, GOF) and remained significant after corrections of multiple comparisons.

### *ESR1* locus variants and association with *TP53* mutation status

We found evidence that multiple *ESR1* locus variants were associated with *TP53* mutation status. In our discovery study, ten variants showed a trend towards association (*P* value <1×10^-4^) for one or more of the three *TP53* mutation phenotypes. From breast cancer GWAS, multiple variants near *ESR1* have been associated with breast cancer of all subtypes as well as ER-tumors [76,78–81]. Some variants at the *ESR1* locus have been reported to exhibit ancestry-specific association with breast cancer risk [81–83].

For example, *ESR1* variant rs140068132 which is thought to have originated in Indigenous Americans, is protective for breast cancer risk [83]. In ER-breast tumors, *TP53* and *ESR1* mutations tend to be mutually exclusive [84]. This may be due in part to the regulatory relationship between TP53 and ESR1. Mutant *TP53* is correlated with lower *ESR1* gene expression which is thought to be due in part to TP53 binding to the *ESR1* promoter to activate expression [85]. Mutant *TP53* tumors have lower estrogen response signatures compared to *TP53* wildtype tumors which may be caused by both decreased transcriptional activation of *ESR1* by mutant TP53 and increased levels of *ESR1*-targeting miRNAs [84]. These studies suggest the possibility that mutation of *TP53* may be an early event that promotes lineage towards ER-breast tumors; it is possible that variants at the *ESR1* locus may enhance or reverse this association. Further functional studies are warranted to understand the connection between *ESR1* variants, *TP53* mutational status, and breast cancer subtypes.

### Variants associated with *TP53* mutation status

Other variants of interest that showed evidence of association with *TP53* included rs17103093 which was associated with any *TP53* mutation phenotype (discovery OR 1.54, *P* value 3.3×10^-5^ and multi-ancestry validation OR 1.4, *P* value 0.03) and maps to an intron of *TACC2*. This variant did not show evidence of association with *TP53* mutations in the MyBrCa study. *TACC2* encodes one of three homologous coiled-coiled proteins (TACC1, TACC2 and TACC3); it shows increased expression in higher grade breast tumors and is associated with local recurrence and reduced survival [86,87]. Variants at the *TACC2* locus are associated with risk of low-grade breast cancer, overall breast cancer, and epithelial ovarian cancer [81,88,89]. Two variants, rs6703393 and rs6890674, showed consistent direction of association for *TP53* GOF mutations in the discovery analyses (OR 0.79, *P* value 7.5×10-5) and the MyBrCa study (OR 0.28, *P* value 0.003) but had no evidence of association in the multi-ancestry validation study (*P* values 0.99 and 0.83 respectively). rs6709393 maps near the *RAB17* gene which encodes for a small GTPase associated with invasion [90]. rs6890674 is located in the 3’UTR of *CD180*, an orphan toll-like receptor that is expressed on B cells and is involved in inflammatory and autoimmune diseases [91].

rs10931697, located near *SLC39A10*, showed opposite effects by study, as it demonstrated increased risk for GOF *TP53* status for the C allele in the discovery study (OR 1.82, *P* value 2.79×10^-5^) but protective effects in the multi-ancestry validation (OR 0.53, *P* value 0.001) and MyBrCa studies (OR 0.72, *P* value 0.09). Further studies are needed to determine if findings represent a false association or if there are ancestry-related differences driving the findings. SLC39A10 is a zinc transporter whose expression is associated with breast cancer metastasis and increased migration *in vitro* [92]. Another variant showing inconsistent directionality of association with a *TP53* mutation phenotype included rs4767726 that maps within a long non-coding RNA (*LNC-RNA 02439*) of which little is known. The C allele of rs4767726 was associated with reduced likelihood of having a *TP53* GOF mutation in the discovery dataset (OR 0.67, *P* value 0.0002) but increased likelihood for a GOF mutation in the multi-ancestry datasets (OR 1.49, *P* value 0.009).

### Germline variants associated with *PIK3CA* mutations and impact on the PI3K/AKT pathway

A variant near *LPAR1*, rs2026801, showed evidence of association in the discovery dataset (*P* value <1×10^-5^) for both any *PIK3CA* mutation and activating *PIK3CA* mutation phenotypes and was the top variant associated with activating *PIK3CA* mutations in the MyBrCA study (G allele OR 0.5, 95% CI 0.38-0.65; *P* value 0.009). However, it showed only a trend towards association in the multi-ancestry analysis (OR 0.88, *P* value 0.16). This variant maps near multiple GWAS hits for hip-to-waist ratio [93] and birth weight [94–96]; both of which are phenotypes associated with increased breast cancer risk [97–99] but has not been associated with risk in women of European, African, or East Asian descent [100]. The nearest gene to rs2026801, *LPAR1,* encodes the lysophosphatidic acid receptor which activates the PI3K/AKT pathway and whose expression is elevated in ER+HER- and lower stage breast cancers [101–103]. In the GTEx database, rs2026801 maps to an expression quantitative trait locus (eQTL) for *LPAR1* in venous blood with the G allele showing lower expression (*P* value 3.93×10^-11^). These data suggest that lower expression of *LPAR1* may provide a cellular environment less likely to require activating *PIK3CA* somatic mutations during tumor development. Further studies are needed to evaluate this hypothesis.

An intronic variant in *KDM4C*, rs10975835, showed evidence of association with *PIK3CA* H1047R/L mutations in the discovery set (OR 1.54, *P* value 1.1×10^-5^) and in the multi-ancestry validation study (OR 1.5, *P* value 0.011), but not in the MyBrCa study (OR 0.92, *P* value 0.66). KDM4C is a lysine demethylase that removes a methylation mark from histones, specifically converting H3K9Me3, a repressive histone mark, to H3K9Me2, a more active mark [104]. Interestingly, a coding variant in *KDM4C*, rs2296067, renders the protein resistant to caspase-3 cleavage and is associated with worse outcomes in breast cancer [105]. *KDM4C* expression is increased in TNBC in which it is thought to be important in chromosomal stability and proliferation [105,106]. In prostate cancer cells, knock-down of *KDM4C* leads to decreased activation of the AKT pathway [107]. KDM4C acts as a lysine demethylase for the TP53 protein leading to decreased TP53 activity and apoptosis [108]. It is unknown if rs10975835 impacts *KDM4C* expression.

Two variants on chromosome 7 (rs194537 and rs9791638) showed suggestive evidence of association with *PIK3CA* p.E545K mutations in the MyBrCa study (unadjusted *P* values 0.042 and 0.047) but not in the multi-ancestry study (*P* values 0.53 and 0.78, respectively). These variants maps near *STEAP2*, a gene in which low expression is associated with poorer outcomes in breast cancer and activation of the PI3K/AKT/mTOR pathway. Conversely, up-regulation of STEAP2 suppresses this pathway [109].

The validation studies included the *MAP3K1*/*SETD9* variants rs252913 and rs331499 reported in the literature to be associated with *PIK3CA* mutation status in ER+ breast tumors, [63]. These variants and others at the *MAP3K1/SETD9* locus have been associated with breast cancer risk in multiple GWAS as well as body size [79,93,110,111]. Neither variant showed evidence of association with *PIK3CA* mutation status in our multi-ancestry validation studies (*P* values 0.54 and 0.37), but both showed suggestive evidence in MyBrCa Study (*P* values 0.09 and 0.14) and the B-CAUSE study (*P* values 0.005 and 0.008) for association with overall *PIK3CA* mutation status. These results should be taken with caution given the very small number of *PIK3CA* mutation carriers in the B-CAUSE study.

Other variants of potential interest included a *MGAT5* intronic variant, rs2442046, and two *SNX13*/*HDAC9* locus variants, rs13230836 and rs6955337. The *MGAT5* variant showed a similar but non-significant effect size for association with p.E542K mutations in the discovery, multi-ancestry validation and MyBrCa studies (ORs 1.86, 1.64, and 1.92). *MGAT5* encodes N-acetylgluocosaminyltransferase V which is a glycosyltransferase important in the synthesis of branched N-glycans, many of which are found on key cancer signaling proteins including EGFR and TGFBR [112,113]. *MGAT5* expression is increased in multiple tumor types; in breast tumors overexpression is associated with metastasis, epithelial to mesenchymal transitions, and cell motility [114]. In mice, MGAT5 and PTEN act antagonistically to regulate PI3K/AKT signaling [115]. rs13230836 and rs6955337 map to chromosome 7 between *SNX13* and *HDAC* and are in LD. They show evidence of association with *PIK3CA* mutations in the discovery (*P* values 7.4×10^-6^ and 3.2×10^-6^) and multi-ancestry validation (*P* values 0.023 and 0.007, respectively) but show non-significance findings in the MyBrCa study. rs6955337 maps within 10kb of rs11560253, a variant associated with gastric cancer risk [116].

However. rs6955337 and rs11560253 are only in LD in European and Japanese population (r^2^>0.8) but not in African or Han Chinese populations (r^2^<0.6). HDAC9 is a class II histone deacetylase whose gene is hypermethylated in HR+ and/or *TP53* wildtype breast cancers [117]; conversely, increased *HDAC9* expression is seen in more aggressive breast cancers and those with poorer prognosis [118,119]. Increased *HDAC9* expression in cell line models is associated with enhanced proliferation, decreased apoptosis and decreased *ESR1* mRNA [118,120].

### Ancestral differences in *TP53* and *PIK3CA* mutation frequencies across cancer types

Associations with genetic ancestry and specific somatic driver mutations have been observed in other cancer types [23,121]. Genetic ancestry is associated with specific somatic driver mutations in *EGFR*, *KRAS* and *STK11* in lung cancer in individuals of Indigenous American ancestry relative to those of EUR or EAS ancestry [122,123]. *TP53* mutations are found at a higher frequency in individuals of AFA relative to individuals of EUR tumors in multiple tumor types (lung, colon, gastric, human papilloma virus negative head and neck), suggesting that genetic background and/or differences in exposures/socio-determinants of health may influence selection of *TP53* somatic mutations [124–127]. *PIK3CA* somatic mutations also show differences by ancestry in different tumor types. For example, *PIK3CA* mutations have been observed at lower frequencies in bladder tumors arising in EAS individuals and in head and neck squamous cell carcinomas from AFA individuals [128,129].

Conversely, *PIK3CA* mutations are more often observed in colorectal tumors from AFA individuals [130]. Variants identified in this study may have utility in explaining *TP53* and *PIK3CA* somatic mutation frequencies arising in different tissues that differ by genetic ancestry. We did not observe any significant ancestry-specific associations after corrections for multiple comparisons, but we were not adequately powered to assess this.

### Study Limitations

There are some limitations to this study. Our discovery analyses were performed in non-Hispanic individuals of EUR, which means that variants enriched in or specific to non-European populations may not have been identified. We were underpowered to determine if our GxM findings were responsible for the observed differences in breast cancer *TP53* and *PIK3CA* mutation frequency for individuals of non-European populations and for variants associated with specific *PIK3CA* mutations (e.g. p.E542K, p.E545K and p.H1047R/L). In our validation study, we did not genotype all variants/loci with *P* values of less than 1×10^-4^ observed in our discovery set, some of which were not included due to low MAF in one or more populations. As such, we may have missed key variants/loci associated with *TP53* or *PIK3CA* mutation status. The source of somatic mutation information used as our phenotype varied widely with some information coming from clinical reports, some from whole genome/whole exome sequencing of tumors, some from targeted sequencing studies, and some from in-house Sanger sequencing studies. Next-generation sequencing is more sensitive than Sanger sequencing for somatic mutations that are present in fewer than 20% of cells or for tumors with a high degree of immune or stromal infiltrate. Our study was based on the premise that *TP53* and *PIK3CA* mutations would be early driver events in tumor development and mutations in these genes should be present in a high proportion of tumor cells. In a previous study, in which we evaluated types of *TP53* mutation by self-reported race and ethnicity, we found no differences in *TP53* mutation frequency across studies by modality of somatic variation detection suggesting that Sanger sequencing is reasonable for mutation detection of early driver events present in a large proportion of cells [14]. Copy number information was not available for a large proportion of tumors; thus, *TP53* mutations due to larger deletions (e.g. chromosome 17p loss) were not included. We expect that a subset of tumors defined as not having a mutation in *TP53* may have had large copy number losses at that locus resulting in the missing of individuals with LOF mutations due to larger deletions.

Across populations, somatic mutations in *TP53* are more common in TNBC and HER2+ tumors; conversely, somatic mutations in *PIK3CA* are much more frequent in ER+ tumors and Luminal breast cancers [2,4]. Even with adjustment based on tumor subtype, it is difficult to sort out the association of the SNV with somatic mutation versus association of the SNV with tumor subtype. Previous studies stratifying by ER- and ER+ tumor status have found ancestry differences in mutation frequency for these genes, but this was not the case for all studies stratifying by tumor subtype [4,8]. Future mechanistic studies are needed to determine if germline variants help drive tumor subtypes that are characterized by certain gene mutations and/or if germline variants impact a cellular context in which a particular mutation is more likely to be selected and the mutation is important for determining tumor subtype.

## CONCLUSIONS

This study provides evidence that germline variants may shape somatic mutation processes or mutation selection of *TP53* and *PIK3CA* in breast tumors. In the future, polygenic risk scores based on findings from this study could identify individuals who are at increased risk of *TP53* or *PIK3CA* mutations should they develop breast cancer which may ultimately inform prevention strategies, such as potential vaccination-based prevention for high-risk individuals more likely to carry a specific somatic mutation. Larger multi-ancestry studies are warranted to confirm the study findings and determine if germline variants explain some of the differences in *TP53* and *PIK3CA* breast cancer mutation frequencies by genetic ancestry. Functional and mechanistic studies are needed to understand the target genes and pathways for variants associated with these mutations in breast tumors.

## DECLARATIONS

### AUTHOR CONTRIBUTIONS

Study conception and design (McElroy, Toland), Experimental Design (Ramroop, Tjader, Toland, Teo, Yao), cohort and sample collection, participant ascertainment and data extraction (Aeilts, Ambrosone, Carpten, Hampel, Huo, Neuhausen, Palmer, Ping, Teo, Toland, Yao, Zhang, Ziv), mutational analysis (Beer, Dauch, Gandhi, Ghanekar, Gillespie, Hu, Liu, Meadows, Pan, Paredes, Ramroop, Sotelo, Tjader, Wei,), Genotyping (Fadda, Pan, Ramroop, Tjader, Toland), and data analysis (Ho, Hu, Liu, Jia, McElroy, Ping, Stevens, Tai, Teo, Wei). All authors provided edits and approved the final draft of manuscript.

## ACKNOWLEDGMENTS

The Stefanie Spielman breast bank and TCC from the OSU CCC Biospecimen Services Shared Resource provided de-identified samples and data for the multi-ancestry validation studies. The Ohio State University GSR performed the targeted genotyping studies and Sanger sequencing of tumor samples. <u>TCGA Data</u>: The results published here are in whole or part based upon data generated by The Cancer Genome Atlas managed by the NCI and NHGRI. Information about TCGA can be found at http://cancergenome.nih.gov. <u>METABRIC:</u> This study makes use of data generated by the Molecular Taxonomy of Breast Cancer International Consortium. Funding for the project was provided by Cancer Research UK and the British Columbia Cancer Agency Branch. <u>TCGA Banerji Study:</u> This work was a collaboration of the Broad Institute in Cambridge, MA and the National Institute of Genomic Medicine (INMEGEN) in Mexico City, Mexico. The work was conducted as part of the Slim Initiative for Genomic Medicine, a project funded by the Carlos Slim Health Institute in Mexico. <u>Nigerian Breast</u> <u>Cancer Data</u>: We are greatly indebted to all the patients who agreed to participate in this study and graciously donated their biological materials. <u>COH Latina Breast Cancer Study</u>: We want to thank the New York Genome Center for the quality of the sequencing and analytic services provided. We also want to thank QIAGEN for their generous donation of PAXgene Tissue Containers and DNA Extraction Kits for this study. This work is dedicated to the memory of the late Mrs. Anne Olorunde (project officer from LASUTH) and Dr. Olayiwola Oluwasola. <u>MyBrCa:</u> The Malaysian Breast Cancer Genetic study thanks all the study participants and all research staff at Cancer Research Malaysia, University Malaya, and Subang Jaya Medical Centre who assisted in recruitment, interviews, and samples/data processing. MyBrCa also thanks the Breast Cancer Association Consortium (BCAC), University of Cambridge and Genome Quebec for their genotyping related work and Caldas Lab and the Core Genomics Facility at the CRUK Cambridge Institute for their sequencing work. <u>AABCG:</u> Data analyses for the AABCG were conducted using the Advanced Computing Center for Research and Education (ACCRE) at Vanderbilt University.

## ETHICS APPROVAL AND CONSENT TO PARTICIPATE

This study was approved by the Ohio State University Cancer Institutional Review Board (IRB) (protocol number 2005C0082). All data and samples were from de-identified individuals who had undergone informed consent for participation in research studies. Patients and sample ids reported in the supplementary information are de-identified (cannot reveal study participant identify) or are not able to be linked to participant identity outside of the researchers on those studies. <u>Nigerian Study:</u> The VOH IRB and the University of Chicago IRB approved study for participants enrolled at their respective sites. <u>COH Latina Study:</u> One hundred and twenty Latina breast-cancer patients seen at City of Hope (COH) in Duarte, California were included in this study. All participants signed a written informed consent approved by the COH Institutional Review Board. MyBrCa was approved by the Independent Ethics Committee, Ramsay Sime Darby Health Care (reference no: 201208.1), and the Medical Ethics Committee, University Malaya Medical Centre (reference no: 842.9).

## DATA AVAILABILITY STATEMENT

The majority of data generated or analyzed during this study are included in this published article (Supplementary Tables S2-S35), in TCGA, dbGAP and/or the following data repositories as listed below. TCGA tumor mutation data and SNV genotyping data are available in dbGAP under accession numbers phs001687.v1.p1, phs000178.v11.p8, and phs002387.v1.p1. METABRIC sequencing data of tumors and SNV genotyping data are available on the European Genome-Phenome archive using accession numbers EGAD0001000164, EGAS00000000083, EGAD00010000158, EGAD00010000266, EGAS00001004518 and EGAD00001006399. The Welcome Trust Sanger Institute data are available in the European Genome-Phenome archive using accessing number EGAS00001001178 and EGAD0010000915. Sequencing data and processed genomic data from the Nigerian breast cancer cases are in dbGAP under study accession number phs001687.v1.p1. Tumor/normal WES and RNAseq data and accompanying phenotypic and clinical/histologic data for the COH Latina Breast Cancer Study are deposited in dbGAP (dbGaP Study Accession: phs003218)[71]. MyBrCa WES and sWGS files are available on the European Genome-phenome Archive under the study accession number EGAS00001004518. Access to controlled patient data will require the approval of the MyBrCa Tumour Genomics Data Access Committee upon request to. Sequence and genotyping data for the Banerji et al. study [72] are available in dbGAP under accession number phs000369.v1.p1. Summary-level statistics genotyping data for the AABCG study are available at GWAS Catalog (accession number: GCST90296719, GCST90296720, GCST90296721, and GCST90296722). B-CAUSE TNBC sequencing data is in the process being deposited into dbGaP with accession number pending.

## REFERENCES

1. 1. The Cancer Genome Atlas Network. Comprehensive molecular portraits of human breast tumours. Nature 2012; 490(7418):61–70. Doi: 10.1038/nature11412.

2. Keenan T, Moy B, Mroz EA, Ross K, Niemierko A, Rocco JW et al. Comparison of the genomic landscape between primary breast cancer in African American versus white women and the association of racial differences with tumor recurrence. J Clin Oncol. 2015; 33(31):3621–3627. doi: 10.1200/JCO.2015.62.2126.

3. Pitt JJ, Riester M, Zheng Y, Yoshimatsu TF, Sanni A, Oluwasola O et al. Characterization of Nigerian breast cancer reveals prevalent homologous recombination deficiency and aggressive molecular features. Nat Commun. 2018; 9(1):4181. doi: 10.1038/s41467-018-06616-0

4. Huo D, Hu H, Rhie SK, Gamazon ER, Cherniack AD, Liu J et al. Comparison of breast cancer molecular features and survival by African and European ancestry in the Cancer Genome Atlas. JAMA Oncol. 2017; 3(12):1654–1662. doi: 10.1001/jamaoncol.2017.0595.

5. Ademuyiwa FO, Tao Y, Luo J, Weilbaecher K, Ma CX. Differences in the mutational landscape of triple-negative breast cancer in African Americans and Caucasians. Breast Cancer Res Treat. 2017; 161(3):491–499. doi: 10.1007/s10549-016-4062-y.

6. Yuan J, Hu Z, Mahal BA, Zhao SD, Kensler KH, Pi J et al. Integrated analysis of genetic ancestry and genomic alterations across cancers. Cancer Cell 2018; 34(4):549–560. doi: 10.1016/j.ccell.2018.08.019.

7. Kamran SC, Xie J, Cheung ATM, Mavura MY, Song H, Palapattu EL et al. Tumor mutations across racial groups in a real-world data registry. JCO Precision Oncology 2021; 5:1654–1658. doi: 10.1200/PO.21.00340.

8. Arora K, Tran TN, Kemel Y, Mehine M, Liu YL, Nandakumar S et al. Genetic ancestry correlates with somatic differences in a real-world clinical cancer sequencing cohort. Cancer Discov. 2022; 12(11):2552–2565. doi: 10.1158/2159-8290.CD-22-0312.

9. Miyashita M, Bell JSK, Wenric S, Karaesmen E, Rhead B, Kase M et al. Molecular profiling of a real-world breast cancer cohort with genetically inferred ancestries reveals actionable tumor biology differences between European ancestry and African ancestry patient populations. Breast Cancer Res 2023; 25(1):58. doi: 10.1186/s13058-023-01627-2.

10. Omilian AR, Wei L, Hong CC, Bandera EV, Liu S, Khoury T et al. Somatic mutations of triple negative breast cancer: a comparison of Black and White women. Breast Cancer Res Treat 2020; 182(2):503–509. doi: 10.1007/s10549-020-05693-4.

11. Yamamoto S, Iwakuma T. Regulators of Oncogenic Mutant TP53 Gain of Function. Cancers (Basel). 2018; 11(1):4. 10.3390/cancers11010004.

12. Alexandrova EM, Mirza SA, Xu S, Schulz-Heddergott R, Marchenko ND, Moll UM. p53 loss-of-heterozygosity is a necessary prerequisite for mutant p53 stabilization and gain-of-function in vivo. Cell Death Dis. 2017; 8(3):e2661. 10.1038/cddis.2017.80.

13. 13. Billant O, Léon A, Le Guellec S, Friocourt G, Blondel M, Voisset C. (2016) The dominant-negative interplay between p53, p63 and p73: A family affair. Oncotarget 2016; 7(43):69549–69564. 10.18632/oncotarget.11774.

14. Pollock NC, Ramroop JR, Hampel H, Troester MA, Conway K, Hu JH et al. Differences in somatic TP53 mutation type in breast tumors by race and receptor status. Breast Cancer Res Treat. 2022; 192(3):639–648. 10.1007/s10549-022-06509-3.

15. Sun X, Xue A, Qi T, Chen D, Shi D, Wu Y et al. Tumor mutational burden is polygenic and genetically associated with complex traits and diseases. Cancer Res 2021; 81(5):1230–1239. doi: 10.1158/0008-5472.CAN-20-3459.

16. Wang S, Pitt JJ, Zheng Y, Yoshimatsu TF, Gao G, Sanni A et al. Germline variants and somatic mutation signature of breast cancer across populations of African and European ancestry in the US and Nigeria. Int J Cancer 2019; 145(12):3321–3333. doi: 10.1002/ijc.32498.

17. Greenblatt MS, Chappuis PO, Bond JP, Hamel N, Foulkes WD. TP53 mutations in breast cancer associated with BRCA1 or BRCA2 germ-line mutations: distinctive spectrum and structural distribution. Cancer Res 2001; 61(10):4092–4097.

18. Manié E, Vincent-Salomon A, Lehmann-Che J, Pierron G, Turpin E, Warcoin M et al. High frequency of TP53 mutation in BRCA1 and sporadic basal-like carcinomas but not in BRCA1 luminal breast tumors. Cancer Res 2009; 69(2):663–671. doi: 10.1158/0008-5472.CAN-08-1560.

19. Holstege H, Joosse SA, van Oostrom CT, Nederlof PM, de Vries A, Jonkers J. High incidence of protein-truncating TP53 mutations in BRCA1-related breast cancer. Cancer Res 2009; 69(8):3625–3633. doi: 10.1158/0008-5472.CAN-08-3426.

20. Natrajan R, Mackay A, Lambros MB, Weigelt B, Wilkerson PM, Manie E et al. A whole-genome massively parallel sequencing analysis of BRCA1 mutant oestrogen receptor-negative and -positive breast cancers. J Pathol. 2012; 227(1):29–41. doi: 10.1002/path.4003.

21. Chen CC, Feng W, Lim PX, Kass EM, Jasin M. Homology-directed repair and the role of BRCA1, BRCA2 and related proteins in genome integrity and cancer. Annu Rev Cancer Biol. 2018; 2:313–336. doi: 10.1146/annurev-cancerbio-030617-050502.

22. Ramroop JR, Gerber MM, Toland AE. Germline variants impact somatic events during tumorigenesis. Trends Genet. 2019; 35(7):515–526. doi: 10.1016/j.tig.2019.04.005.

23. Pereira B, Chin SF, Rueda OM, Vollan HK, Provenzano E, Bardwell HA et al. The somatic mutation profiles of 2,433 breast cancer refines their genomic and transcriptomic landscapes. Nat Commun. 2016; 7:11479, 2016. doi: 10.1038/ncomms11479.PMID: 27161491

24. Nik-Zainal S, Davies H, Staaf J et al. Landscape of somatic mutations in 560 breast cancer whole-genome sequences. Nature 2016; 534(7605): 47–54. doi: 10.1038/nature17676.

25. Tharin Z, Richard C, Derangère V, Ilie A, Arnould L, Ghiringhelli F et al. PIK3CA and PIK3R1 tumor mutational landscape in a pan-cancer patient cohort and its association with pathway activation and treatment efficacy. Sci Rep. 2023; 13(1):4467. doi: 10.1038/s41598-023-31593-w.

26. The TP53 database tp53.isp-cgc.org

27. Petitjean A, Mathe E, Kato S, Ishioka C, Tavtigian SV, Hainaut P et al. Impact of mutant p53 functional properties on TP53 mutational patterns and tumor phenotype: lessons from recent developments in the IARC TP53 database. Hum Mutat. 2007; 28(6):622–629. doi: 10.1002/humu.20495.

28. Giacomelli AO, Yang X, Lintner RE, McFarland JM, Duby M, Kim J et al. Mutational processes shape the landscape of TP53 mutations in human cancer. Nat Genet. 2018; 50(10):1381–1387. doi: 10.1038/s41588-018-0204-y.

29. Kotler E, Shani O, Goldfeld G, Lotan-Pompan M, Tarcic O, Gershoni A et al. A systematic p53 mutation library links differential functional impact to cancer mutation pattern and evolutionary conservation. Mol Cell 2018; 71(5):178–190. doi: 10.1016/j.molcel.2018.06.012.

30. Fortuno C, Lee K, Olivier M, Pesaran T, Mai PL, de Andrade KC et al. Specifications of the ACMG/AMP variant interpretation guidelines for TP53 variants. Hum Mutat. 2021 42(3);223-236. doi: 10.1002/humu.24152.

31. Fortuno C, Pesaran T, Dolinsky J, Yussuf A, McGoldrick K, Tavtigian SV et al. An updated quantitative model to classify missense variants in the TP53 gene: a novel multifactorial strategy. Hum Mutat. 2021; 42(10):1351–1361. doi: 10.1002/humu.24264.

32. Bouaoun L, Sonkin D, Ardin M, Hollstein M, Byrnes G, Zavadil J et al. TP53 variations in human cancers: New lessons from the IARC TP53 database and genomics data. Hum Mutat. 2016; 37(9):865–876. doi: 10.1002/humu.23035.

33. Mizuarai S, Yamanaka K, Kotani H. Mutant p53 induces the GEF-H1 oncogene, a guanine nucleotide exchange factor-H1 for RhoA, resulting in accelerated cell proliferation in tumor cells. Cancer Res. 2006; 66(12):6319–6326. doi: 10.1158/0008-5472.CAN-05-4629.

34. Slovackova J, Grochova D, Navratilova J, Smarda J, Smardova J. Transactivation by temperature-dependent p53 mutants in yeast and human cells. Cell Cycle 2010; 9(11):2141–2148. doi: 10.4161/cc.9.11.11808.

35. Jordan JJ, Inga A, Conway K, Edmiston S, Carey LA, Wu L, Resnick MA. Altered-function p53 missense mutations identified in breast cancers can have subtle effects on transactivation. Mol Cancer Res. 2010; 8(5):701–716. doi: 10.1158/1541-7786.MCR-09-0442.

36. Dittmer D, Pati S, Zambetti G, Chu S, Teresky AK, Moore M et al. Gain of function mutations in p53. Nat Genet. 1993; 4(1):42–46. doi: 10.1038/ng0593-42.

37. Petitjean A, Achatz MI, Borresen-Dale AL, Hainaut P, Olivier M. TP53 mutations in human cancers: functional selection and impact on cancer prognosis and outcomes. Oncogene 2007; 26(15):2157–2165. doi: 10.1038/sj.onc.1210302.

38. Kang HJ, Chun S-M, Kim K-R, Sohn I, Sung CO. Clinical relevance of gain-of-function mutations of p53 in high-grade serous ovarian carcinoma. PLoS One 2013; 8(8):e72609. doi: 10.1371/journal.pone.0072609.

39. Monti P, Campomenosi P, Ciribilli Y, Iannone R, Aprile A, Inga A et al. Characterization of the p53 mutants ability to inhibit p73 beta transactivation using a yeast-based functional assay. Oncogene 2003; 22(34):5252–5260. doi: 10.1038/sj.onc.1206511.

40. Barta JA, Pauley K, Kossenkov AV, McMahon SB. The lung-enriched p53 mutants V157F and R158L/P regulate a gain of function transcriptome in lung cancer. Carcinogenesis 2020: 41(1):67–77. doi: 10.1093/carcin/bgz087.

41. Zerdoumi Y, Lanos R, Raad S, Flaman JM, Bougeard G, Frebourg T, Tournier I. Germline TP53 mutations result into a constitutive defect of p53 DNA binding and transcriptional response to DNA damage. Hum Mol Genet. 2017; 26(14): 2591–2602. doi: 10.1093/hmg/ddx106.

42. Smith PD, Crossland S, Parker G, Osin P, Brooks L, Waller J et al. Novel p53 mutants selected in BRCA-associated tumours which dissociate transformation suppression from other wild-type p53 functions. Oncogene. 1999; 18(15):2451–2459. doi: 10.1038/sj.onc.1202565.

43. Xie X, Lozano G, Siddik ZH. Heterozygous p53 (V172F) mutation in cisplatin-resistant human tumor cells promotes MDM4 recruitment and decreases stability and transactivation of p53. Oncogene. 2016; 35(36):4798–4806. doi: 10.1038/onc.2016.12.

44. Baroni TE, Wang T, Qian H, Dearth LR, Truong LN, Zeng J et al. A global suppressor motif for p53 cancer mutants. Proc Natl Acad Sci USA 2004; 101(14):4930–4935, 2004. doi: 10.1073/pnas.0401162101.

45. Ryan KM, Vousden KH. Characterization of structural p53 mutants which show selective defects in apoptosis but not cell cycle arrest. Mol Cell Biol. 1998; 18(7): 3692–3698. doi: 10.1128/MCB.18.7.3692.

46. Scian MJ, Stagliano KE, Anderson MA, Hassan S, Bowman M, Miles MF, Deb SP, Deb S. Tumor-derived p53 mutants induce NF-kB2 gene expression. Mol Cell Biol. 2005; 25(22): 10097–10110. doi: 10.1128/MCB.25.22.10097-10110.2005.

47. Xu J, Reumers J, Couceiro JR, De Smet F, Gallardo R, Rudyak S et al. Gain of function by mutant p53 by coaggregation with multiple tumor suppressors. Nat Chem Biol. 2011; 7(5):285–295. doi: 10.1038/nchembio.546.

48. West AN, Ribeiro RC, Jenkins J, Rodriguez-Galindo C, Figueiredo BC, Kriwacki R et al. Identification of a novel germ line variant hotspot mutant p53-R175L in pediatric adrenal cortical carcinoma. Cancer Res. 2006; 66(10):5056–5062. doi: 10.1158/0008-5472.CAN-05-4580.

49. Kato S, Han SY, Liu W, Otsuka K, Shibata H, Kanamaru R et al. Understanding the function-structure and function-mutation relationships of p53 tumor suppressor protein by high-resolution missense mutation analysis. Proc Natl Acad Sci USA. 2003; 100(14):8424–8429. doi: 10.1073/pnas.1431692100.

50. Göhler T, Jäger S, Warnecke G, Yasuda H, Kim E, Deppert W. Mutant p53 proteins bind DNA in a DNA structure-selective mode. Nucleic Acids Res. 2005; 33(3):1087–1100. doi: 10.1093/nar/gki252.

51. Friedler A, Veprintsev DB, Hansson LO, Fersht AR. Kinetic instability of p53 core domain mutants: implications for rescue by small molecules. J Biol Chem. 2003; 278:24108–24112. doi: 10.1074/jbc.M302458200.

52. Wu YH, Tsai Chang JH, Cheng YW, Wu TC, Chen CY, Lee H. Xeroderma pigmentosum group C gene expression is predominantly regulated by promoter hypermethylation and contributes to p53 mutation in lunch cancers. Oncogene 2007; 26(33): 4761–4773. doi: 10.1038/sj.onc.1210284.

53. Tsutsumi-Ishii Y, Tadokoro K, Hanaoka F, Tsuchida N. Response of heat shock element within the human HSP70 promoter to mutated p53 genes. Cell Growth Differ. 1995; 6(1):1–8. PMID: 7718482

54. Quinn EA, Maciaszek JL, Pinto EM, Phillips AH, Berdy D, Khandwala M et al. From uncertainty to pathogenicity: clinical and functional interrogation of a rare TP53 in-frame deletion. Cold Spring Harb Mol Case Stud. 2019; 5(4):a003921. doi: 10.1101/mcs.a003921.

55. Dearth LR, Qian H, Wang T, Baroni TE, Zeng J, Chen SW et al. Inactive full-length p53 mutants lacking dominant wild-type p53 inhibition highlight loss of heterozygosity as an important aspect of p53 status in human cancers. Carcinogenesis 2007; 28(2):289–298. 10.1093/carcin/bgl132.

56. Vaughan CA, Frum R, Pearsall I, Singh S, Windle B, Yeudall A et al. Allele specific gain-of-function activity of p53 mutants in lung cancer cells. Biochem Biophys Res Commun. 2012; 429(1):6–10. doi: 10.1016/j.bbrc.2012.09.029.

57. Menendez D, Inga A, Resnick MA. The biological impact of the human master regulator p53 can be altered by mutations that change the spectrum and expression of its target genes. Mol Cell Biol. 2006; 26(6):2297–2308. doi: 10.1128/MCB.26.6.2297-2308.2006.

58. Oh SJ, Im MY. The P53 mutation which abrogates trans-activation while maintaining its growth-suppression activity. Mol Cells. 2000; 10(4):386–391. PMID: 10987134.

59. Lang V, Pallara C, Zabala A, Lobato-Gil S, Lopitz-Otsoa F, Farrás R et al. Tetramerization-defects of p53 result in aberrant ubiquitylation and transcriptional activity. Mol Oncol. 2014; 8(5):1026–1042. doi: 10.1016/j.molonc.2014.04.002.

60. Powers J, Pinto EM, Barnoud T, Leung JC, Martynyuk T, Kossenkov AV et al. A rare TP53 mutation predominant in Ashkenazi Jews confers risk of multiple cancers. Cancer Res. 2020; 80(17):3732–3744. doi: 10.1158/0008-5472.CAN-20-1390.

61. Jeffers JR, Pinto EM, Rehg JE, Clay MR, Wang J, Neale G et al. The common germline Tp53-R337H mutation is hypomorphic and confers incomplete penetrance and late tumor onset in a mouse model. Cancer Res. 2021; 81(9):2442–2456. doi: 10.1158/0008-5472.CAN-20-1750.

62. Benjamini Y, Hochberg Y. Controlling the false discovery rate: a practical and powerful approach to multiple testing. J R Stat Soc Series B 1995; 57(1): 289–300.

63. Puzone R, Pfeffer U. SNP variants at the MAPK1/SETD9 locus 5q11.2 associate with somatic PIK3CA variants in breast cancers. Eur J Hum Genet. 2017; 25(3):384–387. doi: 10.1038/ejhg.2016.179.

64. Smith TR, Liu-Mares W, Van Emburgh BO, Levine EA, Allen GO, Hill JW, Reis IM, Kresty LA, Pegram MD, Miller MS, Hu JJ. Genetic polymorphisms of multiple DNA repair pathways impact age of diagnosis and TP53 mutations in breast cancer. Carcinogenesis 2011; 32(9):1354–1360. doi: 10.1093/carcin/bgr117.

65. Kosoy R, Nassir R, Tian C White PA, Butler LM, Silva G, et al. Ancestry informative marker sets for determining continental origin and admixture proportions in common populations in America. Hum Mutat. 2009;30(1):69–78. doi: 10.1002/humu.20822.

66. Nassir R, Kosoy R, Tian C, White PA, Butler LM, Silva G, et al. An ancestry informative marker set for determining continental origin: validation and extension using human genome diversity panels. BMC Genet. 2009; 10:39. doi: 10.1186/1471-2156-10-39.

67. Kidd JR, Friedlaender FR, Speed WC, Pakstis AJ, De La Vega FM, Kidd KK. Analyses of a set of 128 ancestry informative single-nucleotide polymorphisms in a global set of 119 population samples. Investig Genet 2011;2(1):1. doi: 10.1186/2041-2223-2-1.

68. Wu TD, Reeder J, Lawrence M, Becker G, Brauer MJ. GMAP and GSNAP for Genomics Sequence Alignment: enhancements to speeds, accuracy, and functionality. Methods Mol Bio 206:1418:283–334. doi: 10.1007/978-1-4939-3578-9_15.

69. Dunn T, Berry G, Emig-Agius D, Jiang Y, Lei S, Iyer A et al. Pisces: an accurate and versatile variant caller for somatic and germline next-generation sequencing data. Bioinformatics. 2019; 35(9):1579–1581. doi: 10.1093/bioinformatics/bty849.

70. Cingolani P, Platts A, Wang le L, Coon M, Nguyen T, Wang L et al. A program for annotating and predicting the effects of single nucleotide polymorphisms, SnpEff. Fly (Austin) 2012; 6(2):80–92. doi: 10.4161/fly.19695.

71. Wang K, Li Mingyao, Hakonarson H. ANNOVAR: functional annotation of genetic variants from high-throughput sequencing data. Nucleic Acids Res 2010; 38(16):e164. doi: 10.1093/nar/gkq603.

72. Ding YC, Song H, Adamson AW, Schmolze D, Hu D, Huntsman S et al. Profiling the somatic mutational landscape of breast tumors from Hispanic/Latina women reveals conserved and unique characteristics. Cancer Res. 2023; 83(15):2600–2613. doi: 10.1158/0008-5472.CAN-22-2510.

73. Banerji S, Cibulskis K, Rangel-Escareno C, Brown KK, Carter SL, Frederick AM et al. Sequence analysis of mutations and translocations across breast cancer subtypes. Nature. 2012; 486(7403):405-409. doi: 10.1038/nature11154.

74. Pan JW, Zabidi MMA, Ng PS, Meng MY, Hasan SN, Sandey B et al. The molecular landscape of Asian breast cancers reveals clinically relevant population-specific differences. Nat Commun. 2020; 11(1): 6433. doi: 10.1038/s41467-020-20173-5.

75. Ragu ME, Lim JMC, Ng PS, Yip CH, Rajadurai P, Teo SH et al. TP53 somatic mutations in Asian breast cancer are associated with subtype-specific effects. Breast Cancer Res 2023; 25(1):48. doi: 10.1186/s13058-023-01635-2.

76. Jia G, Ping J, Guo X, Yang Y, Tao R, Li B et al. Genome-wide association analyses of breast cancer in women of African ancestry identify new susceptibility loci and improve risk prediction Nat Genet. Accepted. 2023.

77. Alexander DH, Novembre J, Lange K. Fast model-based estimation of ancestry in unrelated individuals. Genome Res. 2009; 19(9):1655–1664. doi: 10.1101/gr.094052.109.

78. Dunning AM, Michailidou K, Kuchenbaecker KB, Thompson D, French JD, Beesley J et al. Breast cancer risk variants at 6q25 display different phenotype associations and regulate ESR1, RMND1, and CCDC170. Nat Genet. 2016: 48(4);374–386. doi: 10.1038/ng.3521.

79. Michailidou K, Lindström S, Dennis J, Beesley J, Hui S, Kar S et al. Association analysis identifies 65 new breast cancer risk loci. Nature. 2017; 551(7678):92-94. doi: 10.1038/nature24284.

80. Milne RL, Kuchenbaecker KB, Michailidou K, Beesley J, Kar S, Lindström S et al. Identification of ten variants associated with risk of estrogen-receptor-negative breast cancer. Nat Genet. 2017; 49(12):1767–1778. doi: 10.1038/ng.3785.

81. Zheng W, Long J, Gao YT, Li C, Zheng Y, Xiang YB et al. Genome-wide association study identified a new breast cancer susceptibility locus at 6q25.1. Nat Genet. 2009; 41(3):324–328. doi: 10.1038/ng.318.

82. Hoffman J, Fejerman L, Hu D, Huntsman S, Li M, John EM et al. Identification of novel common breast cancer risk variants at the 6q25 locus among Latinas. Breast Cancer Res. 2019; 21(2):3. doi: 10.1186/s13058-018-1085-9.

83. Fejerman L, Ahmadiyeh N, Hu D, Huntsman S, Beckman KB, Caswell JL et al. Genome-wide association study of breast cancer in Latinas identified novel protective variants on 6q25. Nat Commun. 2014; 5:5260. doi: 10.1038/ncomms6260.

84. Li Z, Spoelstra NS, Sikora MJ, Sams SB, Elias A, Richer JK, Lee AV, Oesterreich S. Mutual exclusivity of ESR1 and TP53 mutations in endocrine resistant metastatic breast cancer. NPJ Breast Cancer. 2022; 8(1):62: doi: 10.1038/s41523-022-00426-w.

85. Rasti M, Arabsolghar R, Khatooni Z, Mostafavi-Pour Z. P53 binds to estrogen receptor 1 promoter in human breast cancer cells. Pathol Oncol Res. 2012; 18(2):169–175. doi: 10.1007/s12253-011-9423-6.

86. Cheng S, Douglas-Jones A, Yang X, Mansel RE, Jiang WG. Transforming acidic coiled-coil-containing protein 2 (TACC2) in human breast cancer, expression pattern and clinical/prognostic relevance. Cancer Genomics Proteomics. 2010; 7(2):67–73. PMID: 20335520.

87. Onodera Y, Takagi K, Miki Y, Takayama K, Shibahara Y, Watanabe M et al. TACC2 (transforming acidic coiled-coil protein 2) in breast carcinoma as a potent prognostic predictor associated with cell proliferation. Cancer Med. 2016; 5(8):1973–1982. doi: 10.1002/cam4.736.

88. Purrington KS, Slettedahl S, Bolla MK, Michailidou K, Czene K, Nevanlinna H et al. Genetic variation in mitotic regulatory pathway genes is associated with breast tumor grade. Hum Mol Genet. 2014; 23(22):6034–6046. doi: 10.1093/hmg/ddu300.

89. Lawrenson K, Song F, Hazelett DJ, Kar SP, Tyrer J, Phelan CM et al. Genome-wide association studies identify susceptibility loci for epithelial ovarian cancer in east Asian women. Gynecol Oncol. 2019; 153(2):343–355. doi: 10.1016/j.ygyno.2019.02.023.

90. von Thun A, Birtwistle M, Kalna G, Grindlay J, Strachan D, Kolch W et al. ERK2 drives tumour cell migration in three-dimensional microenvironments by suppressing expression of Rab17 and liprin-B2. J Cell Sci. 2012; 125(Pt 6):1465–1477. doi: 10.1242/jcs.092916.

91. Edwards K, Lydyard PM, Kulikova N, Tsertsvadze T, Volpi EV, Chiorazzi N et al. The role of CD180 in hematological malignancies and inflammatory disorders. Mol Med. 2023; 29(1):97. doi: 10.1186/s10020-023-00682-x.

92. Kagara N, Tanaka N, Noguchi S, Hirano T. Zinc and its transporter ZIP10 are involved in invasive behavior of breast cancer cells. Cancer Sci. 2007; 98(5):692–697. doi: 10.1111/j.1349-7006.2007.00446.x

93. Christakoudi S, Evangelou E, Riboli E, Tsilidis KK. GWAS of allometric body-shape indices in UK Biobank identifies loci suggesting associations with morphogenesis, organogenesis, adrenal cell renewal and cancer. Sci Rep. 2021; 11(1):10688. doi: 10.1038/s41598-021-89176-6.

94. Plotnikov D, Williams C, Guggenheim JA. Association between birth weight and refractive error in adulthood: a Mendelian randomization study. Br J Opthalmol. 2020; 104(2):214–219. doi: 10.1136/bjophthalmol-2018-313640.

95. Yang XL, Zhang SY, Zhang H, Wei XT, Feng GJ, Pei YF et al. Three novel loci for infant head circumference identified by a joint association analysis. Front Genet. 2019; 10:947. doi: 10.3389/fgene.2019.00947.

96. Warrington NM, Beaumont RN, Horikoshi M, Day FR, Helgeland Ø, Laurin C et al. Maternal and fetal genetic effects on birth weight and their relevance to cardio-metabolic risk factors. Nat Genet. 2019; 51(5):804–814. doi: 10.1038/s41588-019-0403-1.

97. Guo W, Key TJ, Reeves GK. Adiposity and breast cancer risk in postmenopausal women: Results from the UK Biobank prospective cohort. Int J Cancer. 2018; 143(5):1037–1046. doi: 10.1002/ijc.31394.

98. Chen C, Chen X, Wu D, Wang H, Wang C, Shen J et al. Association of birth weight with cancer risk: a dose-response meta-analysis and Mendelian randomization study. J Cancer Res Clin Oncol. 2023; 149(7):3925–3935. doi: 10.1007/s00432-022-04171-2.

99. Cao Y, Xia B, Zhang Z, Hu D, Huang X, Yuan J et al. Association of body fat distribution and risk of breast cancer in pre- and postmenopausal women. Obes Facts. 2023; 16(4):356–363. doi: 10.1159/000529834.

100. Jia G, Ping J, Shu X, Yang Y, Cai Q, Kweon SS et al. Genome- and transcriptome-wide association studies of 386,000 Asian and European-ancestry women provide new insights into breast cancer genetics. Am J Hum Genet. 2022; 109(12);2185–2195. doi: 10.1016/j.ajhg.2022.10.011.

101. Liu S, Jiang H, Min L, Ning T, Xu J, Wang T et al. Lysophosphatidic acid mediated PI3K/AKT activation contributed to esophageal squamous cell cancer progression. Carcinogenesis. 2021; 4294):611–620. doi: 10.1093/carcin/bgaa143.

102. Abdelmessih RG, Xu J, Hung FR, Auguste DT. Integration of an LPAR1 antagonist into liposomes enhances their internalization and tumor accumulation in an animal model of human metastatic breast cancer. Mol Pharm. 2023; 20(11):5500–5514. doi: 10.1021/acs.molpharmaceut.3c00348.

103. Benesch MGK, Wu R, Tang X, Brindley DN, Ishikawa T, Takabe K. Lysophosphatidic acid receptor signaling in the human breast cancer tumor microenvironment elicits receptor-dependent effects on tumor progression. Int J Mol Sci 2023; 24(12):9812. doi: 10.3390/ijms24129812.

104. Berry WL, Janknecht R. KDM4/JMJD2 histone demethylases: epigenetic regulators in cancer cells. Cancer Res. 2013; 73(10):2936–2942. doi: 10.1158/0008-5472.CAN-12-4300.

105. Hong Q, Yu S, Yang Y, Liu G, Shao Z. A polymorphism in JMJD2C alters the cleavage of caspase-3 and the prognosis of human breast cancer. Oncotarget. 2014; 5(13):4779–4787. doi: 10.18632/oncotarget.2029.

106. Garcia J, Lizcano F. KDM4C activity modulates cell proliferation and chromosome segregation in triple-negative breast cancer. Breast Cancer (Auckl). 2016; 10:169–175. doi: 10.4137/BCBCR.S40182.

107. Lin CY, Wang BJ, Chen BC, Tseng JC, Jiang SS, Tsai KK et al. Histone demethylase KDM4C stimulates proliferation of prostate cancer cells via activation of AKT and c-Myc. Cancers (Basel). 2019; 11(11):1785. doi: 10.3390/cancers11111785.

108. Lee DH, Kim GW, Yoo J, Lee SW, Jeon YH, Kim SY et al. Histone demethylase KDM4C controls tumorigenesis of glioblastoma by epigenetically regulating p53 and c-Myc. Cell Death Dis 12(1):89. doi: 10.1038/s41419-020-03380-2.

109. Yang Q, Ji G, Li J. STEAP2 is down-regulated in breast cancer tissue and suppresses PI3K/AKT signaling and breast cancer invasion in vitro and in vivo. Cancer Biol Ther. 2020; 21(3):278–291. doi: 10.1080/15384047.2019.1685290.

110. O’Brien KM, Cole SR, Engel LS, Bensen JT, Poole C, Herring AH et al. Breast cancer subtypes and previously established genetic risk factors: a Bayesian approach. Cancer Epidemiol Biomarkers Prev. 2014; 23(1):84–97. doi: 10.1158/1055-9965.EPI-13-0463.

111. Mueller SH, Lai AG, Valkovskaya M, Michailidou K, Bolla MK, Wang Q et al. Aggregation tests identify new gene associations with breast cancer in population with diverse ancestry. Genome Med. 2023; 15(1):7. doi: 10.1186/s13073-022-01152-5.

112. Lajoie P, Partridge EA, Guay G, Goetz JG, Pawling J, Lagana A et al. Plasma membrane domain organization regulates EGFR signaling in tumor cells. J Cell Biol. 2007;179(2):341–356. doi: 10.1083/jcb.200611106.

113. Nagae M, Kizuka Y, Mihara E, Kitago Y, Hanashima S, Ito Y et al. Structure and mechanism of cancer-associated N-acetylglucosaminyltransferase V. Nat Commun. 2018; 9(1):3380. doi: 10.1038/s41467-018-05931-w.

114. Partridge EA, Le Roy C, Di Guglielmo GM, Pawling J, Cheung P, Granovsky M et al. Regulation of cytokine receptors by Golgi N-glycan processing and endocytosis. Science. 2004; 306(5693):120-124. doi: 10.1126/science.1102109.

115. Cheung P, Dennis JW. Mgat5 and Pten interact to regulate cell growth and polarity. Glycobiology. 2007; 17(7):767–773. doi: 10.1093/glycob/cwm037.

116. Jin G, Lv J, Yang M, Wang M, Zhu M, Wang T et al. Genetic risk, incident gastric cancer, and healthy lifestyle: a meta-analysis of genome-wide association studies and prospective cohort study. Lancet Oncol. 2020; 21(10):1378–1386. doi: 10.1016/S1470-2045(20)30460-5.

117. Conway K, Edmiston SN, May R, Kuan PF, Chu H, Bryant C et al. DNA methylation profiling in the Carolina Breast Cancer Study defines cancer subclasses differing in clinicopathologic characteristics and survival. Breast Cancer Res. 2014; 16(5):450. doi: 10.1186/s13058-014-0450-6.

118. Lapierre M, Linares A, Dalvai M, Duraffourd C, Bonnet S, Boulahtouf A et al. Histone deacetylase 9 regulates breast cancer cell proliferation and the response to histone deacetylase inhibitors. Oncotarget. 2016; 7(15):19693–19708. doi: 10.18632/oncotarget.7564.

119. Huang Y, Jian W, Zhao J, Wang G. Overexpression of HDAC9 is associated with poor prognosis and tumor progression of breast cancer in Chinese females. Onco Targets Ther. 2018; 11:2177–2184. doi: 10.2147/OTT.S164583.

120. Linares A, Assou S, Lapierre M, Thouennon E, Duraffourd C, Fromaget C et al. Increased expression of the HDAC9 gene is associated with antiestrogen resistance of breast cancers. Mol Oncol. 2019; 13(7):1534–1547. doi: 10.1002/1878-0261.12505.

121. Henderson BE, Lee NH, Seewaldt V, Shen H. The influence of race and ethnicity on the biology of cancer. Nat Rev Cancer. 2012; 12(9):648–653. doi: 10.1038/nrc3341.

122. Gomez F, Griffith M, Griffith OL. Genetic ancestry correlations with driver mutations suggest complex interactions between somatic and germline variation in cancer. Cancer Discov. 2021; 11(3):534–536. doi: 10.1158/2159-8290.CD-21-0092.

123. Carrot-Zhang J, Soca-Chafre G, Patterson N, Thorner AR, Nag A, Watson J et al. Genetic ancestry contributes to somatic mutations in lung cancer from admixed Latin American populations. Cancer Discov. 2021; 11(3):591–598. doi: 10.1158/2159-8290.CD-20-1165.

124. Kytola V, Topaloglu U, Miller LD, Bitting RL, Goodman MM, D Agostino RB Jr, et al. Mutational landscapes of smoking-related cancers in Caucasians and African Americans: precision oncology perspectives at Wake Forest Baptist Comprehensive Cancer Center. Theranostics.2017; 7(11):2914–2923. doi: 10.7150/thno.20355.

125. van Beek EJAH, Hernandez JM, Goldman DA, Davis JL, McLaughlin K, Ripley RT et al. Rates of TP53 mutation are significantly elevated in African American Patients with Gastric Cancer. Ann Surg Oncol. 2018; 25(7):2027–2033, 2018. doi: 10.1245/s10434-018-6502-x.

126. Cavagna RO, Pinto IA, Escremim de Paula F, Berardinelli GN, Sant’Anna D, Santana I et al. Disruptive and truncating TP53 mutations are associated with African-ancestry and worse prognosis in Brazilian patients with lung adenocarcinoma. Pathobiology. 2023; 90(5):344–355. doi: 10.1159/000530587.

127. Mezghani N, Yao A, Vasilyeva D, Kaplan N, Shackelford A, Yoon A et al. Molecular subtypes of head and neck cancer in patients of African ancestry. Clin Cancer Res. 2023; 29(5):910–920. doi: 10.1158/1078-0432.CCR-22-2258.

128. Peak T, Spiess PE, Li R, Grivas P, Necchi A, Pavlick D et al. Comparative genomic landscape of urothelial carcinoma of the bladder among patients of East and South Asian genomic ancestry. Oncologist. 2023; 28(10):e910–e920. doi: 10.1093/oncolo/oyad120.

129. Mezghani N, Yao A, Vasilyeva D, Kaplan N, Shackelford A, Yoon A et al. Molecular subtypes of head and neck cancer in patients of African ancestry. Clin Cancer Res. 2023; 29(5):910–920. doi: 10.1158/1078-0432.CCR-22-2258.

130. Myer PA, Lee JK, Madison RW, Pradhan K, Newberg JY, Isasi CR et al. The genomics of colorectal cancer in populations with African and European ancestry. Cancer Discov. 2022; 12(5):1282–1293. doi: 10.1158/2159-8290.CD-21-0813.

