## Supplemental Figures for "Association of *ESR1* germline variants with *TP53* somatic variants in breast tumors in a genome-wide study"

**Supplemental Figure 1: Discovery Study Genetic Ancestry Principal Component Analyses**

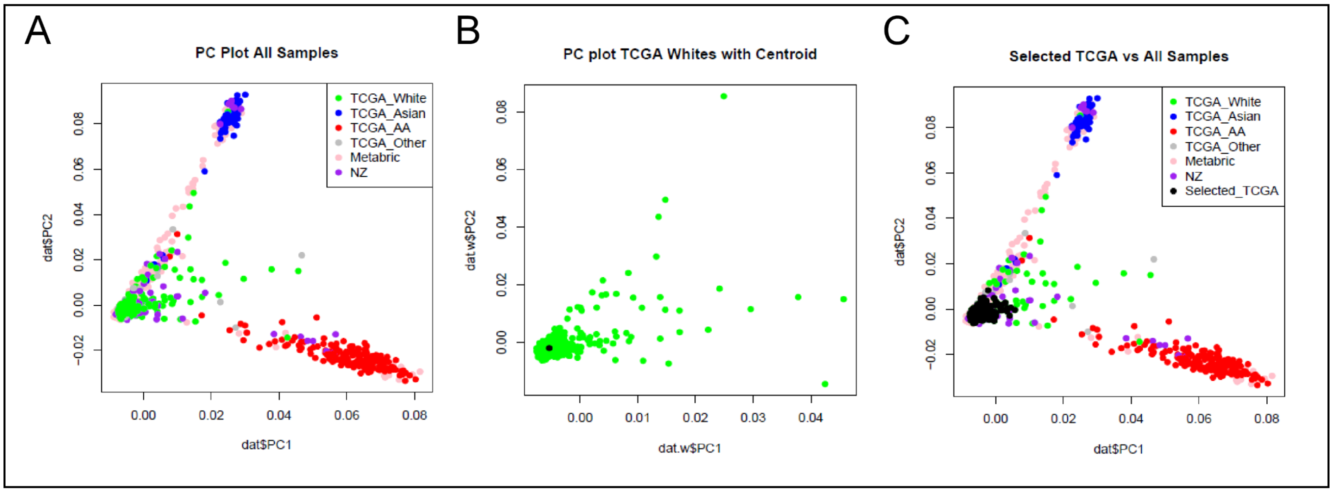


Supplemental Figure 1: Principal component analyses of genetic ancestry are denoted by (A) study and ancestry. (B) PCA of individuals self-identified as white/European race are indicated in green with the centroid indicated as a black dot. (C) Individuals of European genetic ancestry selected for discovery analyses are indicated in black. Individuals in TCGA self-identifying as white/European ancestry TCGA_White (green), black/African ancestry, TCGA_AA (Red), Asian, TCGA_Asian) (Blue) or other race/ethnicity TCGA_Other (Gray). Self-reported race/ethnicity was not included in the METABRIC (pink) or Welcome Trust Sanger (NZ) (purple) studies. PCA, principal component analysis; TCGA, The Cancer Genome Atlas; AA, African Ancestry; METABRIC, Molecular Taxonomy of Breast Cancer International Consortium.

**Supplemental Figure 2: Multi-ancestry Validation Study Genetic Ancestry Principal Components Analyses**

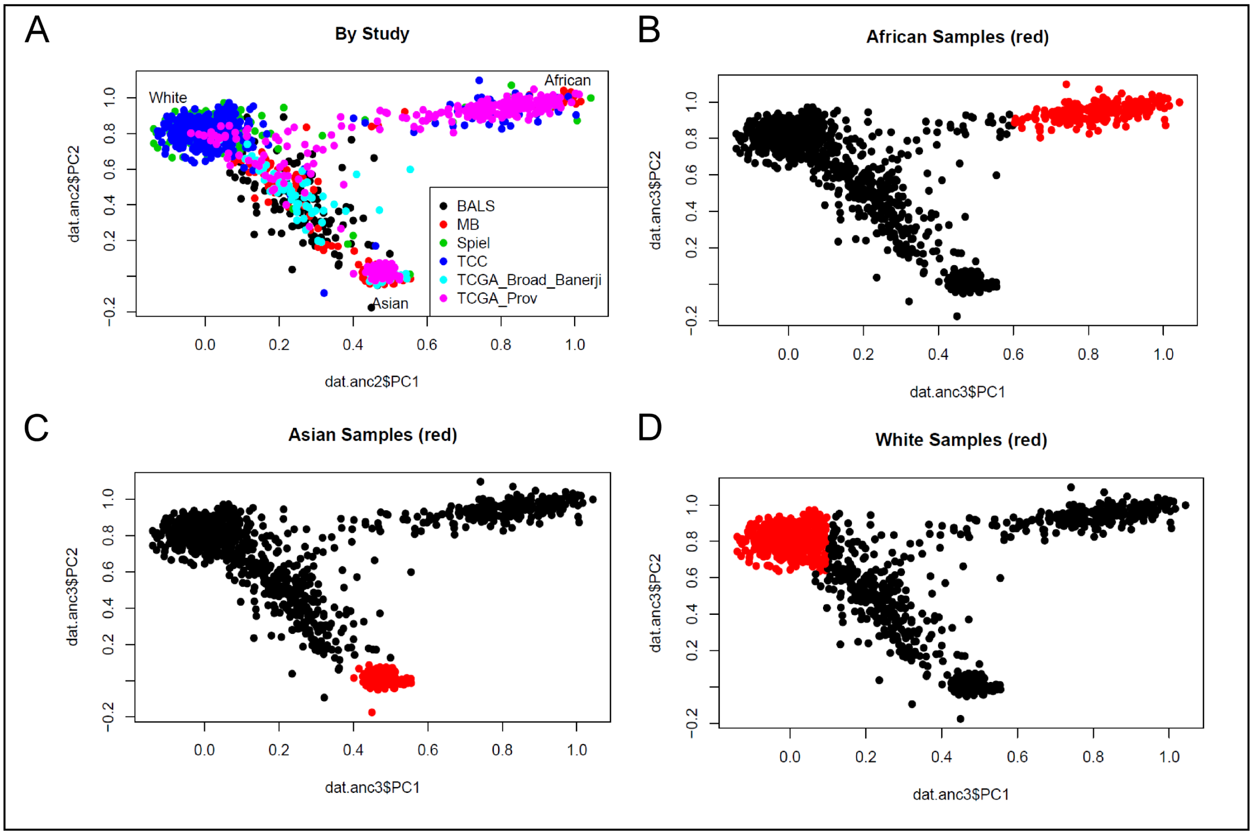


Supplemental Figure 2: Principal component analyses of genetic ancestry are denoted by (A) study cohort or in red by African (B), Asian (C) or European (D) ancestries. Individuals falling between the three major ancestries clusters were assigned as Admixed. BALS, City of Hope Latina Study; MB, METABRIC Molecular Taxonomy of Breast Cancer International Consortium; Spiel, Stefanie Spielman Breast Study; TCC, Total Cancer Care; TCGA_Broad_Banerji, TCGA Banerji study; TCGA_Prov, TCGA Provisional.

**Supplemental Figure 3: TP53 QQ Plots**

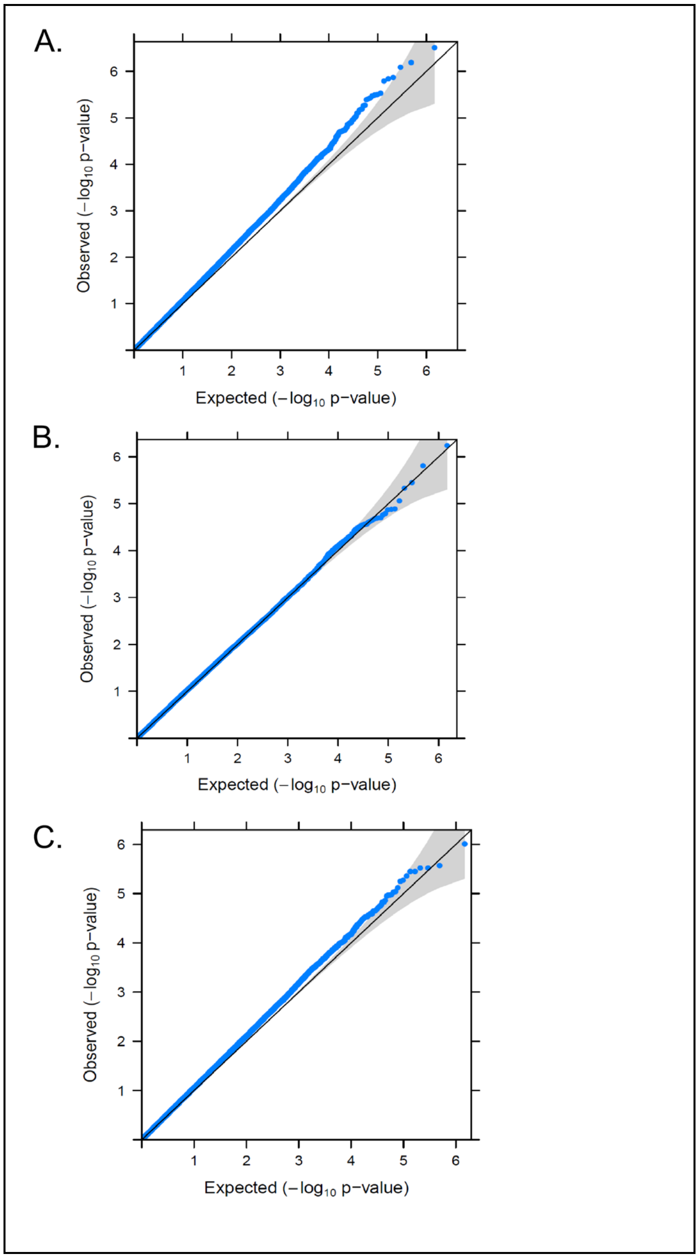


Supplemental Figure 3: *TP53* QQ Plots

QQ plots for the *TP53* GXM are shown for (A) Any *TP53* mutation, (B) GOF *TP53* mutations, and (C) LOF *TP53* mutations. QQ, quantile-quantile; GXM, germline variant by mutation; GOF, gain of function; LOF, loss of function.

**Supplemental Figure 4: PIK3CA QQ Plots**

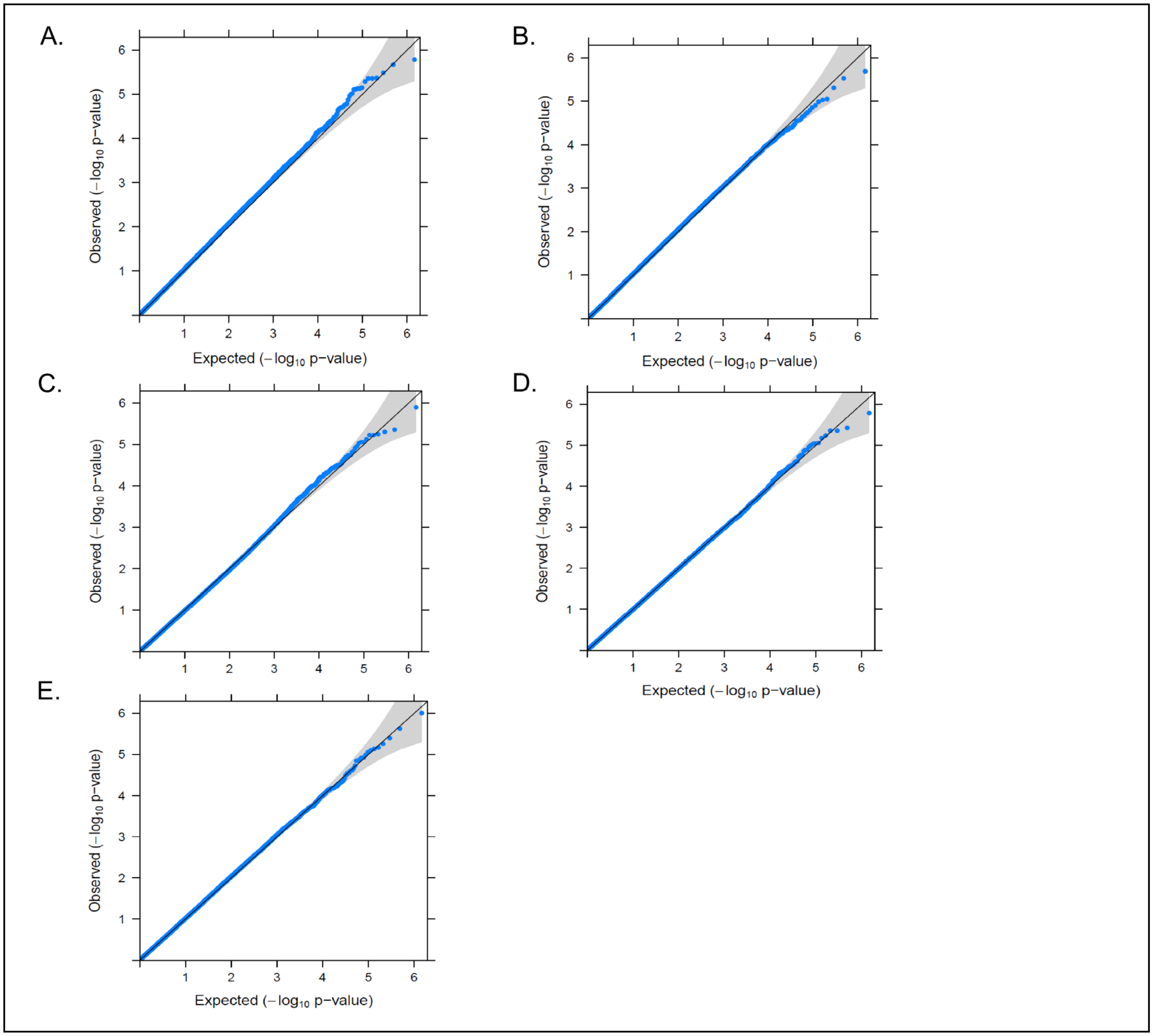


Supplemental Figure 4: *PIK3CA* QQ Plots

QQ plots for the *PIK3CA* GXM are shown for (A) Any *PIK3CA* mutation, (B) Activating/hotspot *PIK3CA* mutations, (C) *PIK3CA* p.E542K mutations, (D) *PIK3CA* p.E545K mutations and (E) *PIK3CA* p.H1047R/L mutations. QQ, quantile-quantile; GXM, germline variant by mutation.
